# Longitudinal AI Models Predict Cancer Adverse Events

**DOI:** 10.64898/2026.10.22.26364879

**Authors:** Chenlian Fu, Jie (Jennifer) Yu, Ruian (Ian) Shi, Michele Waters, Mitchell Pavlak, Andrea Arfe, Sohrab P. Shah, Alexis Battle, Kenneth L. Kehl, Brian M. Bot, US Cancer AI Alliance (CAIA), Nikolaus Schultz, Quaid Morris, Justin Jee

## Abstract

Clinical interventions for cancer treatment toxicities can reduce morbidity, mortality, and healthcare cost, if high risk patients can be identified. We developed SPARC, a transformer-based foundation model, using real-world, longitudinal, routinely collected health measurements, to predict adverse event risk. Pre-training and fine-tuning SPARC on 101,920,379 measurements from 224,047 patients with cancer led to state-of-the-art performance at predicting cancer-related adverse events on (i) held-out data from the development cohort, (ii) a prospective validation cohort from the same center (N_patients_=23,126), (iii) multiple publicly available external datasets (N_patients_=26,012), and (iv) external, multi-hospital cohorts (N_patients_=562,620, 206,840). SPARC generalized to multiple external centers and identified patients most likely to benefit from an intervention (G-CSF). Pooled multi-institution, more than single-institution, fine-tuning improved generalizability. We release SPARC as a community resource. Our results highlight the utility of AI time series models in oncologic management.

## Introduction

Cancer treatment toxicities are a major source of morbidity, healthcare cost, and mortality. Common cytotoxic chemotherapies cause febrile neutropenia ^1^ and acute kidney injury (AKI) in up to a third of patients.^2,3^ Immune checkpoint inhibitors (ICI), now used for a wide range of cancers, can cause serious immune-related adverse events in up to 15% of patients.^4^ Frailer patients at imminent risk of death may benefit from palliative care rather than cancer-directed therapy.^5^ Identifying patients at the highest risk of these adverse events would allow for morbidity-reducing treatment strategies including dose reduction or administration of prophylactic supportive medications.

Quantitative data from longitudinal electronic health records (EHRs), including laboratory values and vital signs, remain underutilized for cancer outcome prediction. Models leveraging continuous clinical time series have shown promise at predicting outcomes in the intensive care unit (ICU),^6,7^ a setting fundamentally different from most cancer care in sampling frequency and treatment setting. EHR data have also been used in forecasting cancer before its formal diagnosis.^8–10^ However, cancer mortality and adverse event prediction has generally relied on static nomograms built on a small number of variables and limited to specific disease and treatment contexts.^11–14^ The full potential of these routinely collected data for cancer side effect prediction remains underexplored.

Given previous findings that deviations from personalized baselines in laboratory values and vital signs are more informative than standard ranges for disease diagnosis,^15^ modeling such patient-specific baseline physiology can inform patient outcome prediction. More generally, the value of leveraging temporal AI models capable of fully capturing longitudinal trends remains unclear in patients undergoing cancer treatment. Here, we used patient-specific clinical time series trajectories for routinely collected quantitative data to train a foundation model to predict cancer treatment-related adverse events and mortality, and evaluated the capacity of the model on key clinical end points.

We hypothesized that transformer-based foundation models trained on routinely collected, longitudinal EHR measurements can better predict cancer treatment-related adverse events and mortality than methods based on static data or single time points. To test this hypothesis, we:

1. Assembled a model development dataset of 224,047 patients with cancer (101,920,379 measurements) from Memorial Sloan Kettering Cancer Center (MSK, New York, NY USA).
2. Developed SPARC (Surveillance of Patient Adverse events using Routine Clinical data), a time-series, transformer-based foundation model pretrained on this institutional dataset.
3. Tested model input and training objectives to optimize SPARC performance for downstream predictive tasks.
4. Tested SPARC and other longitudinal and single-timepoint models to predict clinically actionable treatment-related adverse events and mortality.
5. Assessed model consistency across patient subgroups, identified key predictive features, and demonstrated SPARC’s potential to identify patients most likely to benefit from prophylactic therapy, e.g. granulocyte colony stimulating factor (G-CSF), a prophylactic medication for chemotherapy-induced neutropenia.
6. Further validated these models’ utility in a prospective validation dataset and four external validation datasets spanning patient data from eleven distinct hospitals. We also tested the utility of additional site-specific fine-tuning.

## Results

### Routine clinical time series reflect the personalized nature of cancer progression

We first analyzed a development cohort of 224,047 patients from MSK seen between 2015 and 2021 with at least one measurement for each lab test in the complete blood counts and comprehensive metabolic panels (for demographics see **Table 1**). To characterize the overall structure of these measurements, we computed pairwise Pearson correlations between features across patients, using each patient’s first value per feature. As expected, blood cell counts were correlated according to lineage, with red blood cell and immune related labs clustered respectively. Similarly, measurements for liver enzymes, kidney function markers (BUN and creatinine), and blood pressure were also correlated within each respective module. These modular trends were consistent across cohorts (**Supplemental Figure 1**). As in cohorts of patients without cancer, women had lower red blood cell and creatinine levels, and older patients were more likely to have anemia or reduced renal function (i.e. elevated creatinine and BUN).

In order to assess the patient specificity of different time series features, we then computed the ratio between within-patient, temporal variability and inter-patient variability of each feature. Like others ^15^, we observed (**Supplemental Figure Figure 2**) that intra-patient variability in laboratory values was often greater than inter-patient variability; i.e. individual patients often have unique baselines. As previously identified in healthy patients, hematological lab tests had individualized baselines in patients with cancer. We similarly found individualized baselines in liver function tests, consistent across cohorts. This motivates personalized, time series modeling to better identify individual patient trajectories rather than models based on cohort averages.

### Pretraining encodes clinical states

To create an AI model that can identify meaningful changes in physiology while accounting for individual patient baselines, we pretrained a time series transformer model, SPARC, based on a missingness-aware autoregressive objective. Specifically, SPARC was pre trained to predict lab values at the last time point of a randomly selected patient window. After pretraining on 157K windows containing an average of 10.2 time points and 211.9 measurements per window, SPARC predicted held-out future lab values with a mean squared error (MSE) of 0.423, compared with 0.872 for the feature mean, 0.590 for carrying forward the most recent value, 0.519 for per-feature linear regression on the most recent value, 0.487 for a ridge regression on all most-recent values and their missingness, and 0.451 for a ridge regression on historical lab values. SPARC had lower MSE than each baseline (Wilcoxon signed-rank p < 10⁻⁶ for each comparison; **Supplementary Tables 2–3**). A UMAP of the embedding space confirmed that SPARC’s latent representations, derived from autoregressive pretraining, encode patient future laboratory and vital sign values (**Supplementary Figure 3a**). Embeddings for patients of select cancer types whose abnormalities are captured in routine lab tests also form clusters without labelled supervision (**Supplementary Figure 3b**).

### SPARC prediction tasks and design optimizations

To test whether SPARC and related time series models outperform static time-point models, we trained or fine-tuned multiple models to predict a range of cancer-related adverse events. Because of well-documented concerns about the accuracy of billing codes as ground truth for medical events, we focused on adverse events that were unambiguously encoded in the EHR either through mortality status or quantitative measurements:

A. mortality within 180 days following hospital discharge, which has implications for hospice discharge decisions
B. mortality within 30 days following ICU admission, which may help with triaging of critical care resources
C. anemia, within 30 days of an infusion of a cytotoxic chemotherapy associated with these side effects
D. neutropenia within 30 days of an infusion of a cytotoxic chemotherapy associated with these side effects
E. febrile neutropenia within 30 days of an infusion of a cytotoxic chemotherapy associated with these side effects;
F. AKI within 30 days of either chemotherapy or bisphosphonate infusion;
G. hepatotoxicity within 90 days of infusion of ICI ^16–21^

Details including lab value cutoffs and agents included are provided in **Methods**. For events C-G, each infusion cycle of the relevant medication was considered a separate prediction window in which an event could occur. Only infusions before and up to the first such adverse event per category were included (e.g. if a patient had a neutropenic event during multiple chemotherapy infusion events, only the first infusion with a neutropenia event and any preceding infusions were included as prediction windows).

We optimized SPARC through a series of design studies, varying one component at a time, its feature set, architecture, and loss function. Because these experiments required multiple candidate modalities, we used a harmonized dataset (N=57,510) including tumor genomic profiling with MSK-IMPACT and annotated data ^22,23^ including genetic ancestry, cancer type, organ sites of involvement, socioeconomic status (Yost index), ^24^ functional status (ECOG score) ^25^, smoking status, and less commonly ordered labs. The final architecture was then retrained on the full MSK cohort and validated on held-out, prospective and external datasets.

We evaluated whether adding these less routinely captured modalities improved predictive performance relative to models based on standard laboratory values and vital signs finding that the rarer data modalities did not consistently improve performance (**Supplementary Figure 4a**). As such we restricted subsequent training to laboratory and vital sign measurements. Interestingly, adding medication types as features and training subcohort-specific models also did not improve performance at predicting medication side effects (**Supplementary Figure 4b-c**).

Motivated by the modular structure seen in the correlation analyses (**Supplemental Figure 1**), we found that multi-layer perceptron embeddings grouped by input type (i.e. red blood cell-based, liver-based, etc.) outperformed a single mixed embedding fed all raw inputs in most tasks (**Supplementary Figure 4d**). As a result, models with modular embeddings were used for all downstream analyses.

Most cancer treatment spans multiple cycles. We therefore tested whether combining binary cross-entropy with a time-to-event loss function ^26,27^, where we substitute time to event with number of infusion cycles to side effect onset, would yield performance gains over a purely binarized outcome (see **Methods**, **Supplemental Figure 4e, Supplemental Table 4**). Because of improvements in performance using time-to-event loss, such loss functions were used for medication side effect prediction tasks.

We also investigated whether a more effective representation of time further improves model performance. SPARC maps each measurement’s relative timestamp through periodic functions with learned rates and offsets, then linearly projects the result into an embedding added to the visit’s modular embeddings. Compared with a raw relative timestamp or no time input, this continuous-time encoding matched or improved performance on all seven tasks (**Supplemental Figure 4f**). We thus incorporated this continuous-time encoding into the SPARC architecture.

The SPARC architecture may be trained from scratch to perform prediction tasks without pretraining, though the amount of training data required for this is unclear. In our analysis, pretraining was especially important in a regime with less training data. In all tasks, we see a negative correlation between increasing training data size and the benefit pretraining provides prior to supervised training (p<0.05 in hospital mortality, ICU mortality, anemia, **Supplementary Figure 5**).

This initial design phase informed our final SPARC model (**Figure 1**). The final model uses a modular design built on parsimonious, routinely collected clinical time series features: complete blood count tests (e.g., red blood cell count, hemoglobin concentration), comprehensive metabolic panel tests spanning kidney function (e.g., creatinine), liver function (e.g., ALT), and electrolytes (e.g., sodium), along with routine vital signs (e.g., heart rate). The model undergoes autoregressive pretraining followed by task-specific finetuning, using Cox losses for the adverse event tasks.

**Figure 1.**
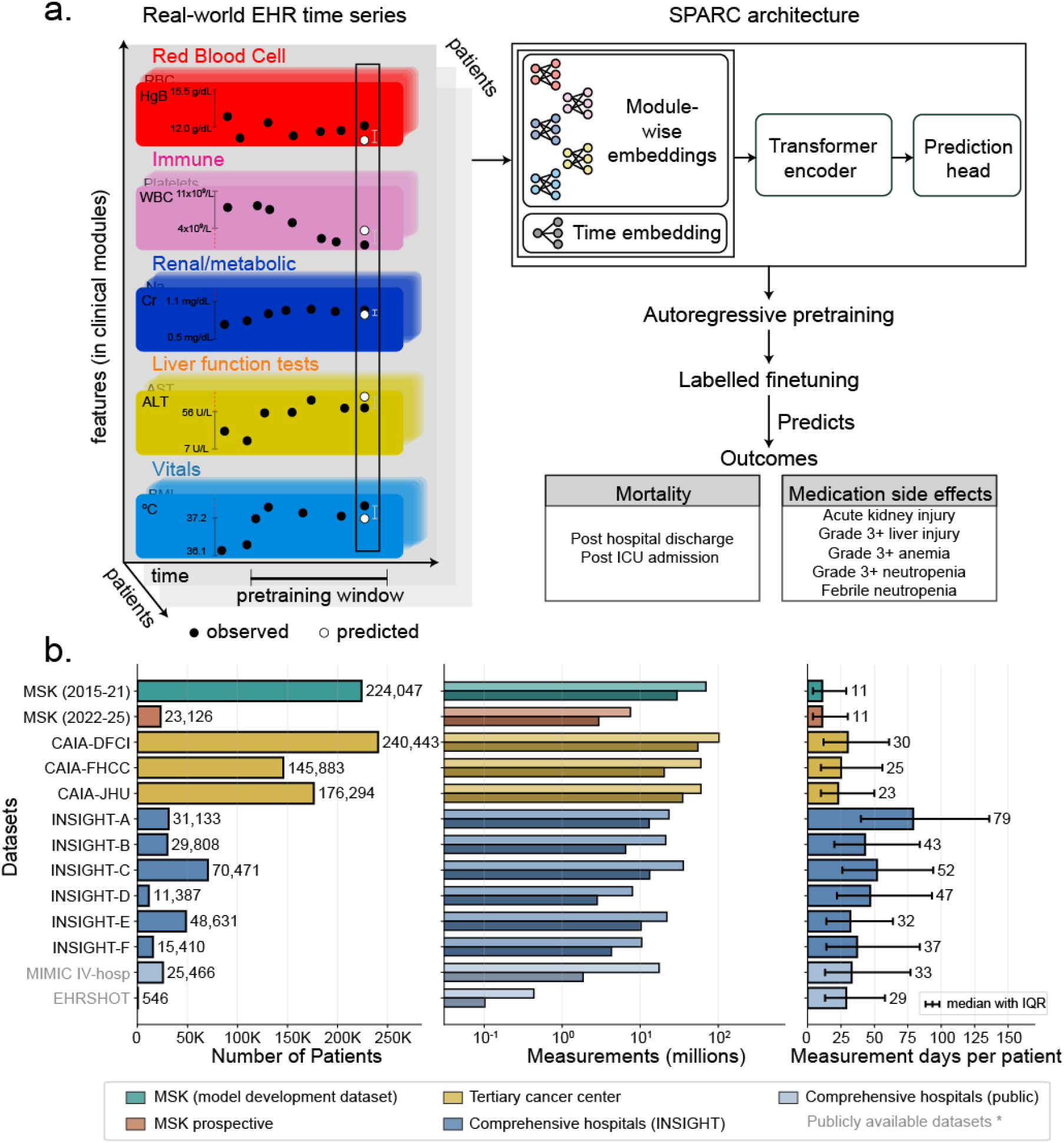
Study overview. **(a)** SPARC schematics. SPARC uses the correlated feature modules and time series in its architecture, is pretrained using an autoregressive reconstruction loss, and is fine-tuned to predict mortality and treatment induced adverse events. **(b)** Datasets used to develop and validate SPARC. SPARC is internally developed at Memorial Sloan Kettering Cancer Center and validated using an internal prospective dataset and 11 external datasets, with varying number of patients, measurements, and number of measurement days per patient. Dataset demographics are in Supplemental Table 1.

### Benchmarking models for cancer adverse event prediction

We tested the performance of SPARC as well as several other AI architectures for either time series or static time point data for predicting cancer adverse events (**Figure 2, Supplemental Table 5**). In a held-out test set, SPARC outperformed or matched all other models at predicting both mortality tasks as well as AKI, anemia, liver toxicity, and neutropenia. TabPFN, a foundation model pretrained on large-volume tabular data ^28^, performed best at predicting the rare febrile neutropenia side effect. Other time series models, including both neural network and tree-based models, did not outperform SPARC or TabPFN at any tasks. In addition, single time point models generally did not outperform time series models. Together, these results highlight the relative utility of pretrained transformer architectures for cancer adverse event prediction tasks using clinical time series data.

**Figure 2.**
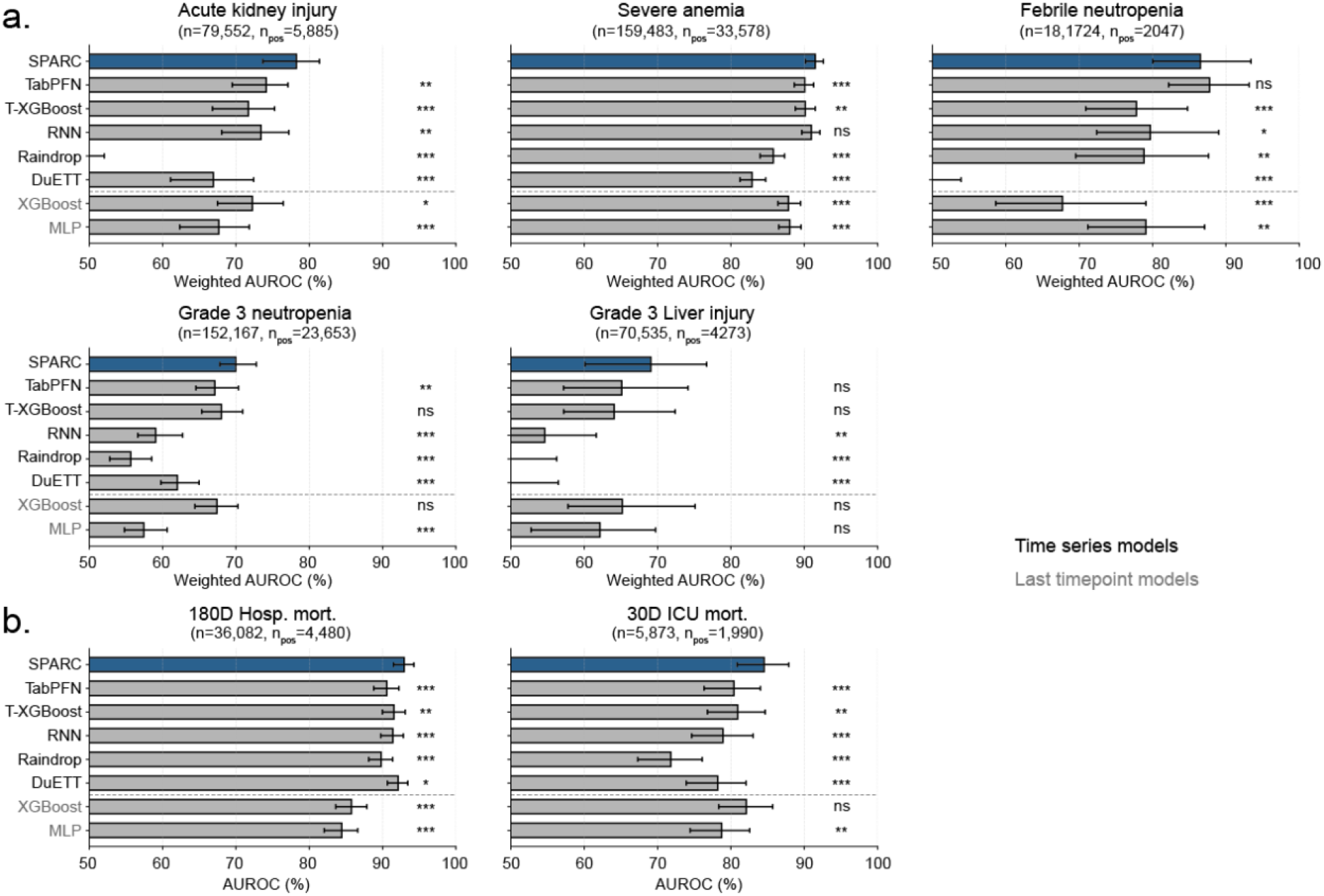
SPARC performance benchmarking. Comparing SPARC model performance with other time series (y axis labels in black) and single/last timepoint benchmarks (y axis labels in grey) in a held-out test set of the MSK dataset, for (a) medication side effect prediction tasks and (b) mortality tasks; *: p<0.05, **: p<0.01, ***: p<0.001 (two-sided paired bootstrap for SPARC vs. one benchmark at a time, 2,000 resamples).

We further compared SPARC performance in AKI, anemia, febrile neutropenia, and anemia with nomograms from literature ^12–14^. As we lacked standard inputs for the nomogram or used different endpoint definitions, we retrained nomograms on our cohort using overlapping features for fairness of comparison. SPARC significantly outperformed both the original and retrained nomograms in anemia and febrile neutropenia prediction and matched the performance of nomograms for neutropenia prediction (**Figure 3a**). As the number of AKI events was small, it was challenging to compare SPARC with nomogram in AKI predictions, although SPARC performed significantly better than random chance, whereas the nomogram did not.

**Figure 3.**
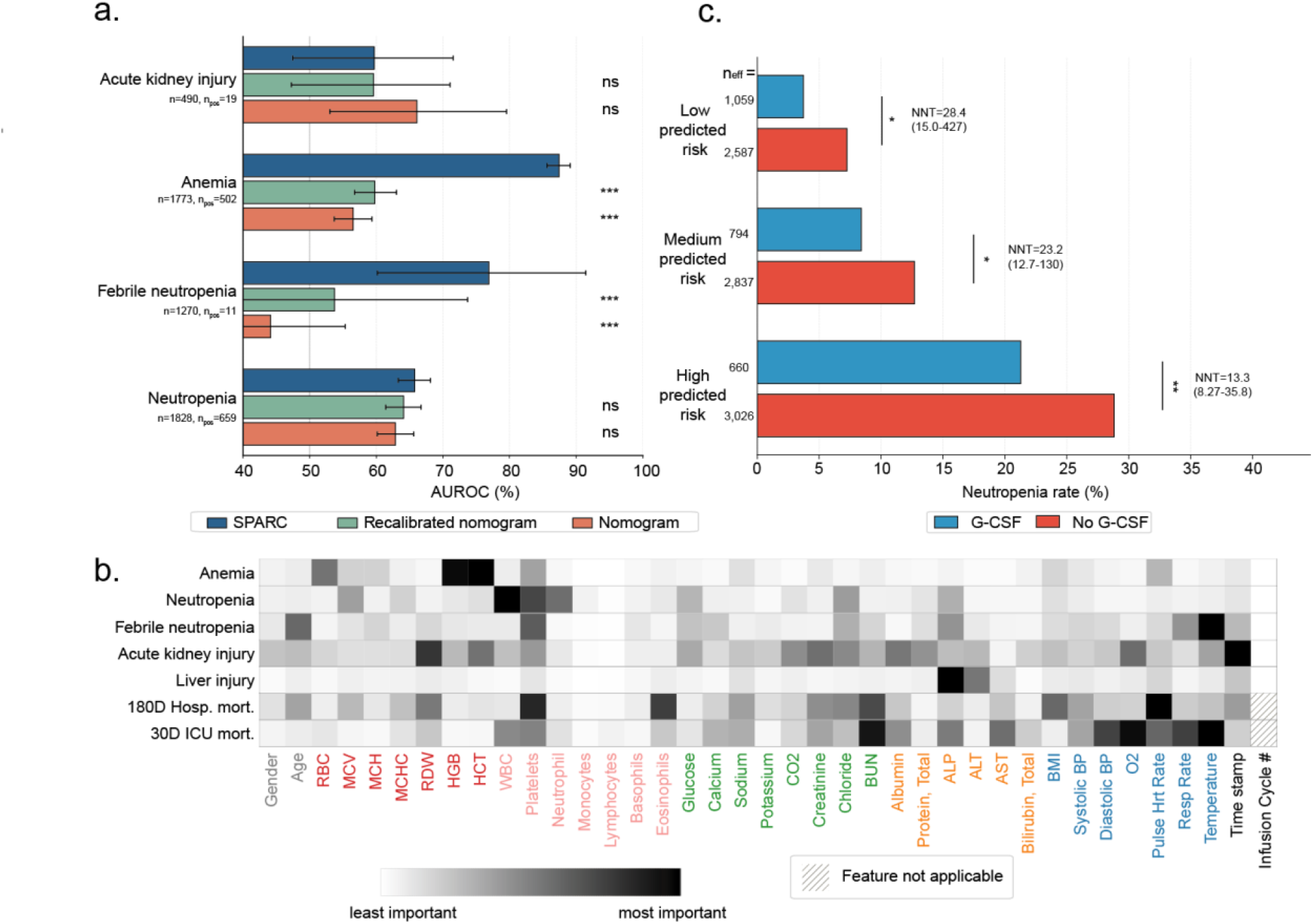
Clinical applications and interpretations of SPARC. **(a)** Comparison of SPARC to existing clinical nomograms for comparable endpoints, see Methods for included nomograms and subcohort definition, and see Supplemental Materials for endpoint definitions (two-sided DeLong test, ***: p<0.001). **(b)** Feature importance analyses of SPARC for each task using LIME, with absolute feature importance averaged over timepoints (unaveraged versions showing all timepoints are in **Supplemental Figure 6**). **(c)** SPARC-predicted neutropenia risk identifies patients most likely to benefit from prophylactic G-CSF. Within tertiles of predicted risk, treated and untreated infusions were balanced by inverse probability of treatment weighting (IPTW); n_eff_, effective sample size. NNT, number needed to treat (1/absolute risk reduction); 95% CIs and p-values from a patient-level bootstrap (*: p < 0.05, **: p < 0.01). IPTW was chosen over propensity score matching (PSM), whose results depended on the number of matched controls but were consistent with IPTW **(Supplementary Figure 8a**); positivity assumption for conducting IPTW and PSM was satisfied **(Supplementary Figure 8b).**

### Model interpretability and subset analyses

We quantified the relative importance of input features to SPARC’s predictions using LIME (**Figure 3b, Supplemental Figures 6**), ^29^ averaging attributions across patients. For the high-level summary in **Figure 3b**, we averaged the magnitude of these attributions over the patient’s history, weighting each time point by its overall contribution. As expected, red cell indices (hematocrit, hemoglobin, and red blood cell count) contributed most to anemia prediction, white blood cell and platelet counts contributed most to neutropenia prediction, and liver function tests (alkaline phosphatase, ALT and AST) contributed most to liver toxicity prediction. Febrile neutropenia prediction was driven chiefly by temperature, platelet count and age. Interestingly, the red cell distribution width (RDW) was the single most important laboratory predictor of AKI. Time relative to the last included measurement was the most important feature for AKI and among the most important for liver toxicity, but contributed less to anemia, neutropenia and 30D ICU mortality prediction. Blood urea nitrogen (BUN) was among the top predictors of both 180D hospital mortality and 30D ICU mortality, whereas ICU mortality prediction relied more heavily on vital signs (temperature, oxygen saturation, respiratory rate and blood pressure). We also observed that feature missingness in CBC differential tests which are less frequently prescribed contributed more to model prediction than the other lab tests. Temporally, the cumulative importance of a patient’s history outweighed that of the single most recent measurement for six of seven tasks, most markedly for 180D hospital mortality, 30D ICU mortality and febrile neutropenia, with anemia being the exception (**Supplemental Figures 6b)**. This demonstrates that SPARC integrates longitudinal clinical information rather than relying on a single most recent snapshot and does so in a task-dependent manner. SPARC thus leverages both diverse clinical and temporal features to accurately predict adverse events.

Each of these prediction tasks span heterogeneous groups of patients. To better understand the predictive power of these models, we performed subset analyses of treatment-related adverse events stratified by treatment regimen. In these analyses, SPARC retained high accuracy in a majority of cases, although performance was heterogeneous in some tasks (**Supplementary Figure 7, Supplemental Table 6**). AKI was most accurately predicted in patients receiving biphosphonates (AUROC 0.87); performance was weaker for patients receiving cisplatin (AUROC 0.64). We performed further stratified analyses based on cancer type. In these analyses, SPARC achieve robust performances in cancer types with large amounts of training data (breast, prostate cancers) as well as sarcoma and lymphoma.

### SPARC supports G-CSF prophylaxis decisions

We sought to assess SPARC’s ability to identify patients who may derive significant benefit from timely prescription of prophylaxis mitigating treatment side effects. We performed Inverse probability of treatment weighting (IPTW) matched analysis on the held-out test cohort to test whether SPARC’s predicted risk strata were associated with the number needed to treat (NNT) with granulocyte colony-stimulating factor (G-CSF) to avoid neutropenia events. Patients who received G-CSF prophylaxis had lower observed neutropenia event rates in the medium and high risk, but not low risk strata. Furthermore, NNT decreased across increasing predicted risk (**Figure 3c, Supplemental Figure 8**). These results demonstrate that SPARC can identify high-risk patients in whom G-CSF prophylaxis is most useful, supporting its use to target prophylactic interventions.

### SPARC generalizes in prospective and external validations

We sought to test the reliability and generalizability of SPARC through validation on orthogonal cohorts, including 1. A prospective validation cohort including 23,126 patients from Memorial Sloan Kettering Cancer Center after data lock for SPARC training, 2. Patients with cancer from the publicly available MIMIC-IV dataset ^30^ (Beth Israel Deaconess Medical Center ICU patients, Boston MA, N=25,466), 3. Patients with cancer from the publicly available EHRSHOT dataset ^31^ (Stanford University Medical Center, Stanford CA, N=546), 4. The CAIA consortium, which includes four NCI-designated cancer centers (total N=562,620), including Johns Hopkins Hospital (Baltimore, MD), Dana-Farber Cancer Institute (Boston, MA), and Fred Hutchinson Cancer Center (Seattle, WA), along with MSK (excluded from this analysis), 5. The INSIGHT dataset ^32^, a multi-hospital consortium including six comprehensive hospitals across New York City, NY and Houston, TX (total N=206,840).

The demographics of these cohorts is presented in **Supplementary Table 1**. More women received care at CAIA tertiary cancer centers compared to comprehensive hospitals at the INSIGHT dataset due a high incidence of breast cancer in the former. Tertiary cancer centers in CAIA included more non-Hispanic White and Asian patients, whereas more Hispanic and Black patients were treated at INSIGHT hospitals.

In the prospective validation cohort, the only task where the performance consistently degraded by more than 5% in AUROC was febrile neutropenia, possibly reflecting the small number of events and changes in patient case mix over time (**Figure 4a**). These results confirm SPARC’s validity across time despite changes in cancer treatment paradigms and temporally associated environmental factors.

**Figure 4.**
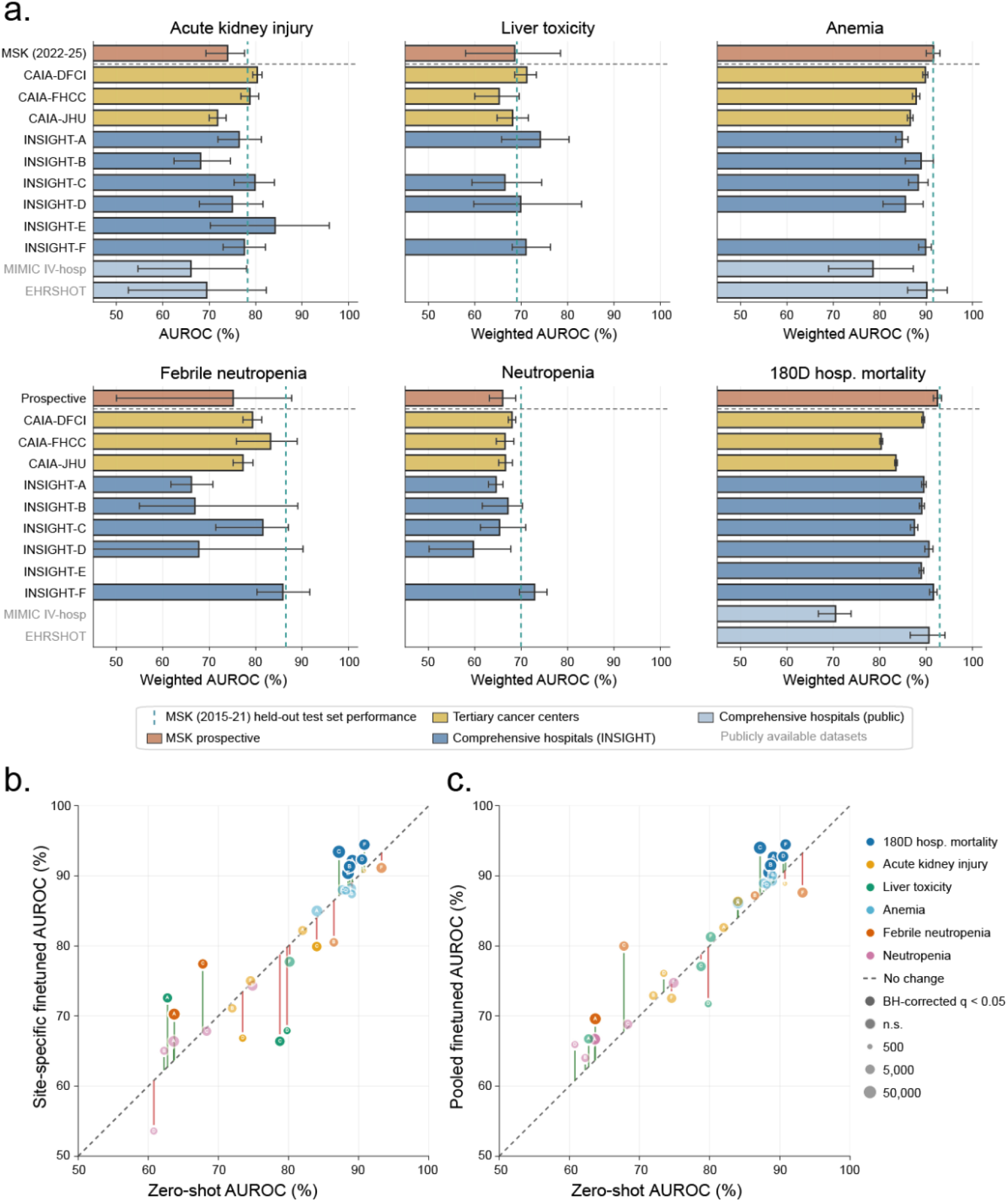
SPARC external validation. We evaluated SPARC performance in an MSK prospective dataset, three CAIA tertiary cancer centers, INSIGHT hospitals, and two publicly available datasets (a) in zero-shot settings (note INSIGHT-B did not have sufficient patients and/or events to be evaluated for liver toxicity; INSIGHT-E did not have sufficient patients and/or events to be evaluated for liver toxicity, anemia, febrile neutropenia, and neutropenia; MIMIC IV-hosp and EHRSHOT did not have sufficient **patients** and/or events not for liver toxicity, febrile neutropenia, and neutropenia) (b) when finetuned and then evaluated using site-specific data at the INSIGHT hospitals, and (c) when finetuned using pooled training data across 6 INSIGHT hospitals and then evaluated using site-specific data. The bubble sizes represent the total number of patients available for a task at an INSIGHT hospital.

We first assessed SPARC zero-shot at the three external cancer centers in the CAIA consortium. Febrile neutropenia was the only task that consistently degraded relative to the MSK held-out set (maximum ΔAUROC ∼10%), again likely reflecting limited training data, lower precision in evaluation metrics for rare events, and institutional differences. For all other tasks, results remained close to, and occasionally exceeded, those at MSK, suggesting the model generalizes to other cancer centers.

We next validated the model at six hospitals in the INSIGHT consortium (anonymized as A–F). Hospital E lacked enough eligible patients to evaluate liver toxicity, febrile neutropenia, and neutropenia. Zero-shot results held up for higher-prevalence tasks, including 180-day mortality, anemia, AKI, and liver toxicity, with AKI and liver toxicity slightly outperforming MSK at hospitals A and E, respectively. Consistent with the CAIA findings, febrile neutropenia and neutropenia proved harder to predict, in line with the greater sensitivity of low-prevalence endpoints to cross-institutional shift.

Finally, we tested SPARC on cancer patients from two public datasets spanning comparable time scales, MIMIC-IV-hosp and EHRSHOT. Results in EHRSHOT were comparable to MSK, whereas MIMIC-IV-hosp showed weaker performance, possibly because its inclusion criteria (ICU or emergency department admission) create patient populations that differ notably from the training population.

### Fine-tuning on external data

Motivated by the demographic shift and performance loss observed in the INSIGHT data, we investigated whether locally fine-tuning MSK-trained SPARC models further improved performance (**Figure 4b)**. We fine-tuned MSK-trained SPARC models in two ways: (i) using only training data from each individual INSIGHT hospital and validating using a held out dataset in that hospital, and (ii) pooled fine-tuning using data from all INSIGHT hospitals and evaluating on held-out data from a single hospital site. Interestingly, pooled fine tuning improved on the zero-shot baseline in more task-site pairs than site-specific fine-tuning (24/30 vs 15/30; discordance 10 vs 1; McNemar exact p = 0.0117). We further found that, by titrating the amount of training data from the pooled training set for model training, performance on most tasks plateaued with one-eighth of the pooled data, whereas liver injury continued to improve with more data, with smaller gains for anemia and mortality (**Supplemental Figure 9**). These findings underscore the value of aggregating data across sites rather than fine-tuning on any single site in isolation, with the amount of pooled data needed varying by outcome.

## Discussion

We have shown that routinely collected EHR time series measurements can predict cancer outcomes and treatment side effects. Pretraining was helpful in many tasks, but not all, and primarily in low data regimes. Integrative time series modeling performed nearly uniformly better than single time points, or nomograms. Without further optimizations, SPARC generalized to other cancer hospital settings, and for many prediction tasks, to non-cancer center hospitals. Adding more training data from other hospitals only improved SPARC’s performance for more challenging tasks, like liver toxicity; for many tasks, SPARC has already achieved maximum performance in contrast to scaling law observations in other domains.^33^

Our results have direct implications for precision supportive care. Patients at highest SPARC-predicted risk of neutropenia were most likely to benefit from G-CSF administration, directly motivating the use of SPARC in interventional, randomized studies of G-CSF administration.

Our study has limitations. The variable performance of SPARC with cancer types and treatment agents may be due to either the inherent difficulty of these tasks or inadequate data. Selection bias can guide which patients receive which medications, with ramifications for prediction; for example, AKI was hard to predict in patients following cisplatin chemotherapy possibly because clinicians are less inclined to prescribe cisplatin to patients with underlying renal dysfunction. We only used SPARC for outcomes clearly defined by laboratory results, further work should consider outcomes derived from clinical notes. For the outcomes studied here, we incorporated additional modalities and found no substantial added predictive values. However, modalities including comorbidities and imaging have proven important for predicting other outcomes, such as treatment response and cancer recurrence ^34–38^. Future efforts can expand these modalities to broaden the range of predictable outcomes. Although we validated SPARC models using large, geographically and demographically diverse cohorts, all these patients were treated at academic medical centers in large US cities. Laboratory testing patterns may differ in community and non-US hospitals due to resource constraints, insurance coverage, and demographic differences.

We present SPARC as a community resource, and we hope that our results support future work in translating AI modeling into further personalization of care.

## Methods

### Patients

The following cohorts were analyzed:

1. Two cohorts of MSK patients for primary analysis, utilizing the parsimonious feature set, as described in *Data Elements*: i) a discovery set including patients from 01/01/2016-12/31/2021, and ii) a prospective validation dataset including patients followed from 01/01/2022-02/01/2026.
2. A multimodal dataset including 57,510 patients from MSK who underwent MSK-IMPACT sequencing. Only patients with at least one set of laboratory and vital sign tests were included. This cohort was used to conduct ablation studies and model design experiments, using the full feature set, as described in the *Data Elements* section.
3. Anonymized datasets from the US Cancer AI Alliance (CAIA) consortium included the following hospital sites: Dana-Farber Cancer Institute, Fred Hutchinson Cancer Center, Johns Hopkins University Medical Center. CAIA patient data included the parsimonious feature set, as described in *Data Elements*.
4. Anonymized datasets including the parsimonious feature set from the INSIGHT-Clinical Research Network (INSIGHT-CRN) were obtained from the following hospital sites: New York University School of Medicine, Albert Einstein College of Medicine (Montefiore Medical Center), Weill Cornell Medicine, Houston Methodist, Columbia University Irving Medical Center, and the Icahn School of Medicine at Mount Sinai. The cohort was restricted to patients aged 21–80 years, of either sex, with qualifying encounters between January 1, 2018 and January 1, 2025.
5. Public datasets included in the analyses entail the cancer subset of the MIMIC-IV hosp and EHRSHOT datasets. MIMIC-IV captures predominantly hospital-based care, with sparse outpatient history and no outpatient chemotherapy administration data, and EHRSHOT is a smaller cohort without ICU stay data. Both datasets were anonymized by applying per-patient date shifting for de-identification prior to its public release. Both datasets are governed by their respective data use agreements and were obtained from PhysioNet under credentialed access. Patients with cancer were selected with ICD-9-CM and ICD-10-CM billing codes for malignant neoplasm.

This study was approved by the Institutional Review Board at MSK (IRB# 25-241), which waived the need for informed consent given the minimal risk posed to patients by the study.

### Data elements

The following data elements were considered for model features during model design. The parsimonious features were included in training and testing models across all datasets. Genomic and additional features were only consistently available for the 57,510-patient, harmonized cohort, which were used in our ablation studies.

#### Laboratory test and vital signs time series features

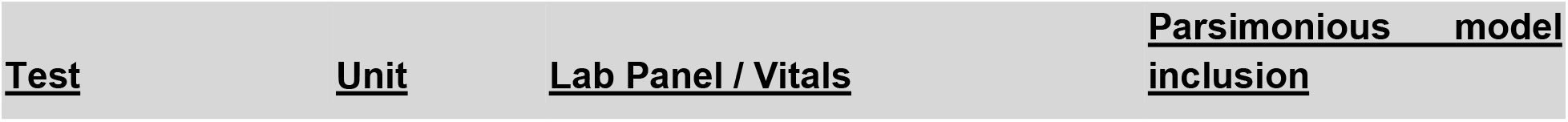
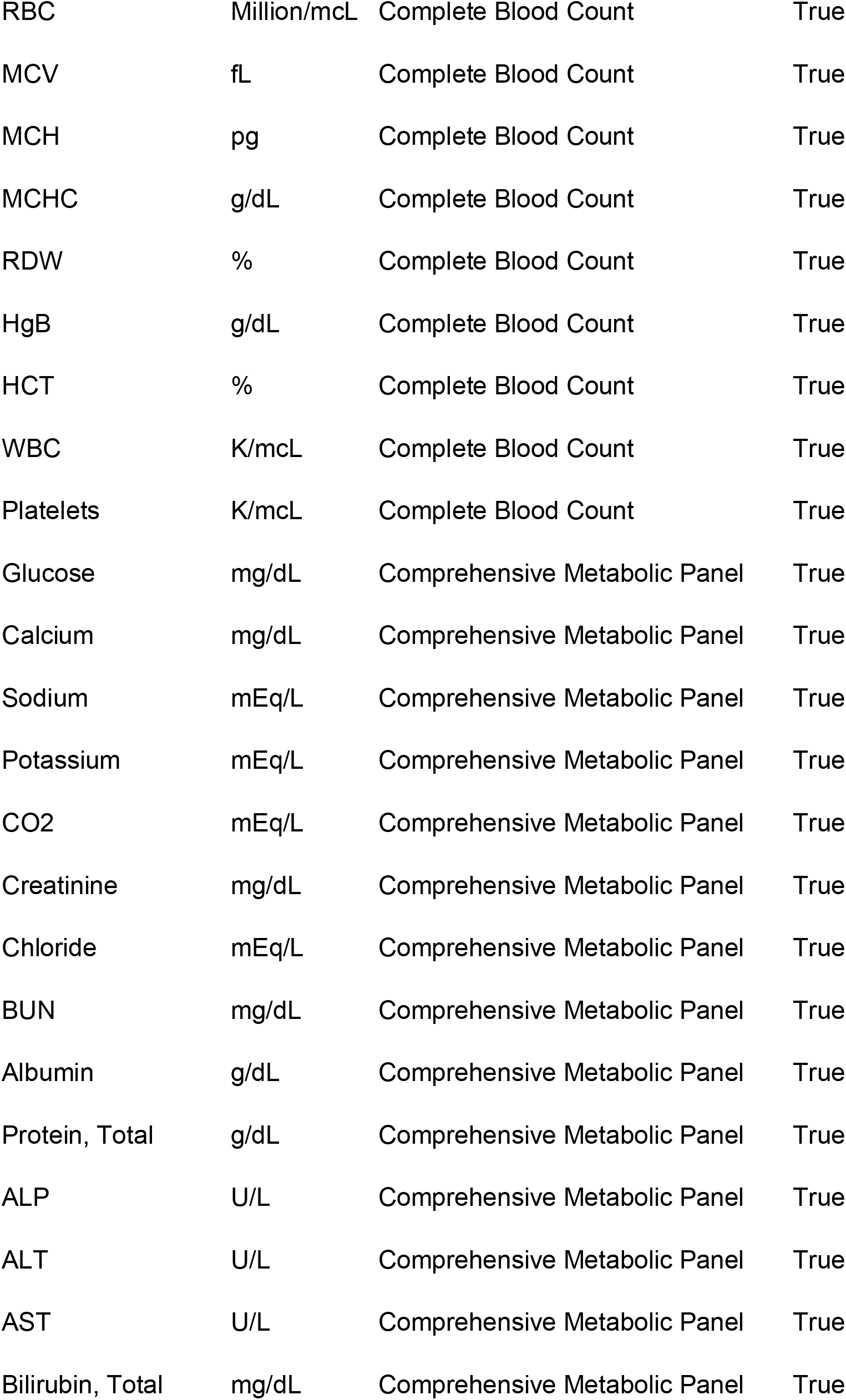
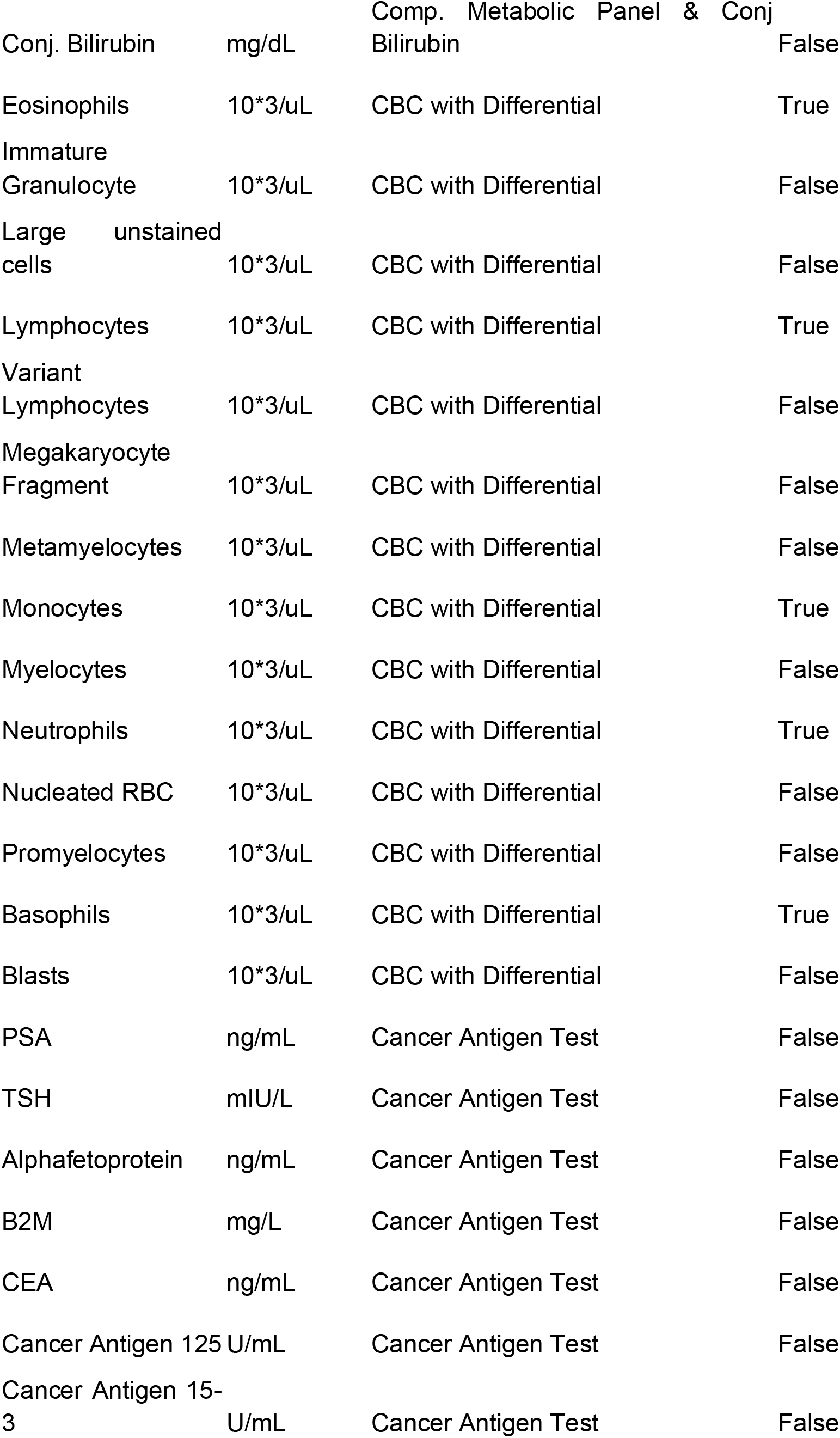
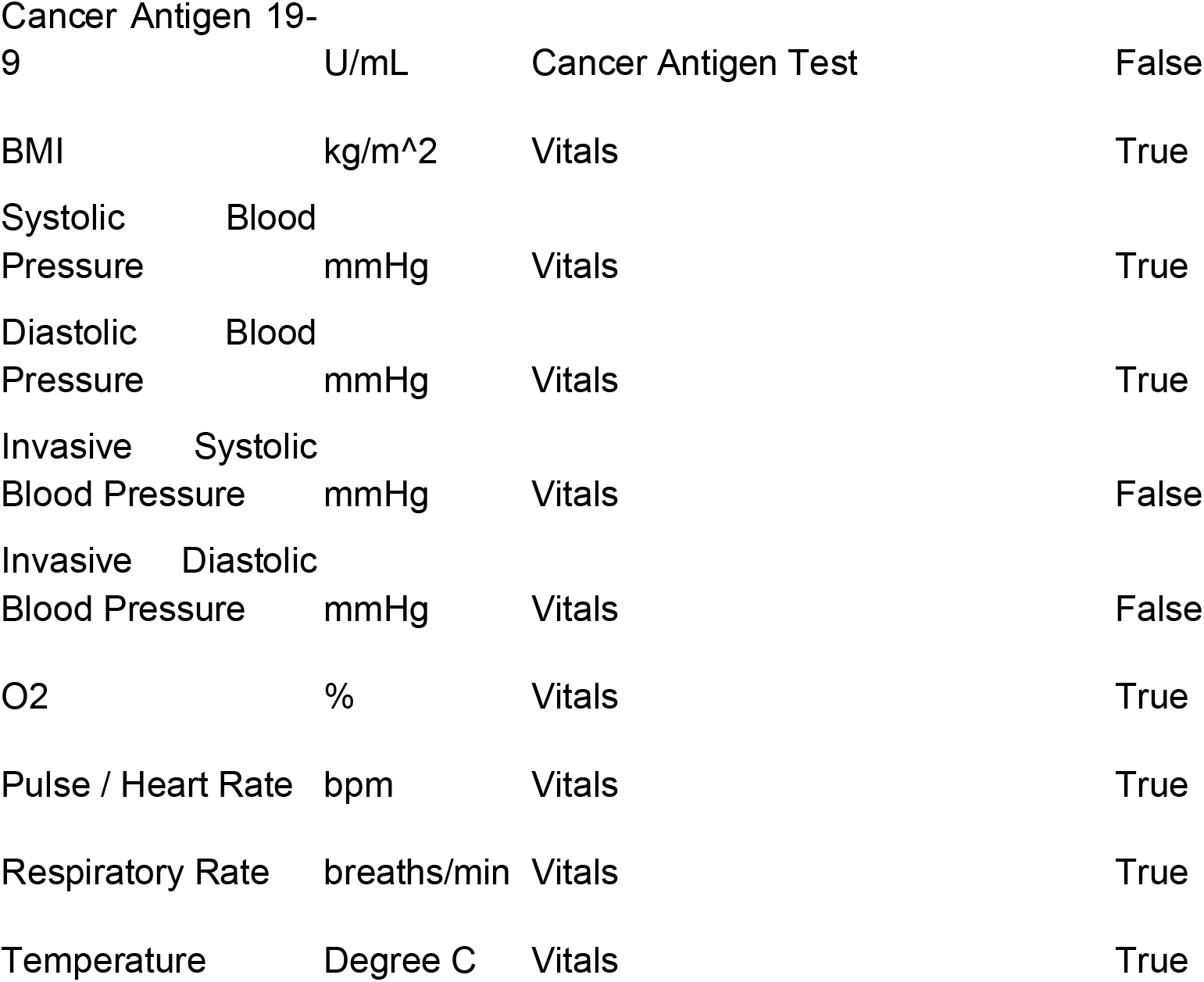

For the INSIGHT, CAIA, and MSK dataset, we included only patients who have at least one measurement of every parsimonious lab or vital signs features except for the CBC differential tests to improve data quality and density, while for MIMIC IV-hosp and EHRSHOT, a patient was included as long as they have ICD 9/10 cancer codes as well as one such test due to limited cohort size.

#### Genomic features

Static genomic features were derived from the MSK-IMPACT targeted tumor sequencing cohort, as described previously^39^ for the harmonized ablation cohort. Each sequenced sample was annotated for the presence of nonsynonymous, truncating and OncoKB-annotated oncogenic mutations in each panel gene; gene- and allele-level cancer hotspot mutations; focal amplifications and deep deletions; arm-level copy-number gains and losses; gene fusions; pathway-level alterations; mutational signatures inferred from trinucleotide substitution contexts; and continuous measures of mutational burden, including tumor mutational burden, fraction of genome altered and microsatellite instability score. To allow samples sequenced on the 341-, 410-, 468- and 505-gene panel versions to be represented consistently, features were restricted to genes on the IMPACT341 panel, which is common to all versions, yielding 5,044 genomic features per sample. Sample metadata and demographic covariates were excluded from this feature block, as they are represented by separate model modules, and any remaining missing values were set to zero. Patients with more than one sequenced sample were represented by their most recently accessioned sample.

#### Additional features

Sex was one-hot encoded with male as the reference level (1 feature). Age at first sequencing was binned into 5-year intervals and passed as a single ordinal value (1 feature). These two features are included in the parsimonious feature set.

Each patient in the harmonized ablation cohort carries a set of additional static features as available from MSKCC cBioPortal repository and the MSK-CHORD dataset ^40^, including the patient-specific Yost index, an area-level socioeconomic measure passed as a continuous value imputed to the cohort median, ranging 1-100; ECOG performance status as a single ordinal value ranging 0-4, with unrecorded status set to 0; smoking status, one-hot encoded over three classes (former/current, never, unknown) with unknown as the reference level (2 features); and primary cancer type, one-hot encoded over 34 categories with “others” as the reference level (33 features), including non-small cell lung, colorectal, breast, prostate, pancreatic, endometrial, ovarian, bladder, esophagogastric, hepatobiliary, head and neck, thyroid, cervical, appendiceal, salivary gland, bone, and small cell lung cancers, renal cell carcinoma, cancer of unknown primary, glioma, melanoma, non-melanoma skin cancer, mesothelioma, soft tissue and uterine sarcoma, gastrointestinal stromal tumor, germ cell tumor, leukemia, mature B-cell and mature T- and NK-cell neoplasms, myeloproliferative neoplasms, myelodysplastic syndromes, and myelodysplastic workup; one-hot encoded genetic ancestry (6 categories: EUR, ADM, EAS, AFR, SAS, NAM); and one-hot encoded site of metastatic disease (10 categories), including adrenal glands, bone, CNS/brain, intra-abdominal, liver, lung, lymph nodes, pleura, reproductive organs, and other.

### Description of prediction tasks

The fine tuning tasks for SPARC are defined as follows:

- 180D hospital mortality: binary classification following the latest available hospital discharge for each given patient, as motivated by the 6-month mortality risk estimate as required by Medicare to reimburse hospice care ^41^. Patients whose last contact was prior to 180 days post discharge and remained alive at that date were excluded.
- 30D ICU mortality: binary classification following the latest available ICU admission for each given patient, to assess the benefit of ICU admission to the patient given only information prior to admission. Patients whose last contact was prior to 30 days post ICU admission and remained alive were excluded.
- Grade 3 anemia: binary classification of whether a patient’s hemoglobin falls below 8 g/dL per CTCAE definition^42^, within 30 days following an infusion (or before the next infusion, whichever comes first) of a treatment regimen including one or more of: docetaxel, sacituzumab govitecan, cisplatin, topotecan, irinotecan, irinotecan liposomal, cyclophosphamide + doxorubicin + vincristine (in combination), bendamustine, carboplatin, etoposide, cabazitaxel. The qualifying medications are selected based on risk of hematological adverse events per NCCN guidelines ^43^.
- Grade 3 neutropenia: binary classification of whether a patient’s absolute neutrophil count falls below 1,000 cells/µL per CTCAE definition^42^, within 30 days following an infusion (or before the next infusion, whichever comes first) of a regimen including one or more of the same medications for grade 3 anemia.
- Febrile neutropenia: binary classification of whether a patient’s absolute neutrophil count falls below 1,000 cells/µL together with a temperature ≥ 38.0 °C measured within 2 days of each other, within 30 days following an infusion (or before the next infusion, whichever comes first) of a regimen including one or more of the same medications for grade 3 anemia.
- Acute kidney injury: binary classification of whether a patient meets KDIGO serum-creatinine criteria for AKI (stage ≥ 1), a rise of ≥ 0.3 mg/dL within 48 hours, or ≥ 1.5X a rolling 7-day baseline (most recent prior value, floored at 0.5 mg/dL ^44^, within 30 days following an infusion (or before the next infusion) of a regimen including one or more of: cisplatin, ifosfamide, pemetrexed, methotrexate, pamidronate, zoledronic acid. The qualifying medications are selected per Onconephrology Core Curriculum 2023^45^.
- Grade 3 liver toxicity: binary classification of whether a patient experiences either bilirubin > 5 mg/dL, or AST / ALT / ALP exceeding 5× baseline per CTCAE definition^42^, within 90 days following an infusion (or before the next infusion) of a regimen including one or more of: ipilimumab, atezolizumab, nivolumab, pembrolizumab, avelumab, cemiplimab, dostarlimab. The qualifying medications are selected as they increase risk of hematological adverse events per ASCO irAE guidelines^46^.

A patient’s infusions after they have a positive event were excluded from further model training or evaluation.

### Cohort-level feature correlation and coefficient of variation analyses

Within each cohort, we characterized the inter-feature and time-dependent properties of laboratory, vital sign, and demographic features.

To assess inter-feature relationships, we computed pairwise Spearman rank correlation coefficients using each patient’s first available value for each feature and visualized the resulting correlation matrices.

To assess time-dependent variability, we computed a CV (coefficient of variation) ratio for each feature using an approach consistent with Foy et al.^15^ The CV is defined as the ratio of the standard deviation to the mean of a given distribution. The inter-patient CV was the CV of all measurements of a feature pooled across patients. To make intra-patient variability independent of follow-up length, the intra-patient CV was computed within 60-day windows and sliding in 30-day steps. For each patient, we took the median CV across windows containing at least three measurements of the feature, and then the median across patients with at least one such window.

The CV ratio is then intra-patient CV divided by the inter-patient CV, where a lower value indicates that a feature’s variability is driven more by within-patient fluctuation over time than by differences across patients.

### SPARC Pretraining

SPARC has a predefined maximum context window of k timepoints (default *k* = 20) and is pretrained using a missing- and time-aware autoregressive objective. All longitudinal features are first z-score normalized per feature across all patients. For each patient with m timepoints (dates) with at least one measurement (*m* ≥ 2), we sample a consecutive window of length a, where a is drawn from [2, min(*m*, 1 + *k*)]. SPARC learns to use the first (*k* − 1) timepoints in the window to predict the last timepoint’s normalized feature values, with mean squared error loss computed over non-missing features at the last timepoint. Timepoints within the context window that are unfilled (*k* − *a* + 1 remaining slots) are handled via padding, and missing features within sampled timepoints are assigned a sentinel value of −10 to distinguish them from z-score normalized non-missing values. Multiple non-overlapping windows may be sampled sequentially from the same patient’s record; to prevent overrepresentation of patients with long clinical histories, sampling is capped at three windows per patient.

### SPARC setup and finetuning

The default, optimized SPARC has the following designs:

- A parsimonious feature set: gender, 5-year binned age, and the parsimonious lab and vital signs features
- Module-specific feed-forward networks prior to transformer encoder: RBC related (HGB, HCT, RBC, MCV, MCH, MCHC, RDW), immune-related (WBC, Neut, Mono, Lymph, Baso, Eos, Platelets), renal and metabolic (Creatinine, BUN, Sodium, Potassium, Chloride, CO2, Calcium, Glucose), hepatic (ALT, AST, ALP, Bilirubin Total, Albumin, Protein Total), routine vitals (BMI, Systolic BP, Diastolic BP, O2, Pulse, Resp Rate, Temperature), age (5-year binned age), gender.
- Missingness for each time series feature is an explicit input, where each module’s feature value projection receives the concatenation of its feature values and their missingness indicators. In addition, each module token is multiplied by the fraction of that module’s constituent features actually observed at that visit.
- Continuous-time encoding: elapsed time is encoded with learned Fourier features of the relative number dates with respect to the last lab/vital date to accommodate irregular inter-visit spacing and variable numbers of visits. 256 frequencies and 256 phases are initialized on a log scale (as in sinusoidal positional encoding) and learned during training; the resulting sine and cosine features are projected through a single linear layer to 512 dimensions and added to the embedding prior to transformer.

10-fold cross validation with one-fold held-out testing was used: data was split randomly into 11 equally sized folds, with one held out for testing; in each of 10 rounds of cross validation, one out of ten non-heldout folds was used for validation and the rest for training. The SPARC model is first pretrained, and then, for each of the 7 tasks, finetuned using the pretrained model serving as weight initialization, both using the 10-fold cross validation set up. In held-out test set evaluation or evaluation using external data, SPARC predictions were obtained by averaging the predicted probabilities of the 10 resulting finetuned models.

### Titration experiments

We sought to further investigate the benefits of pretraining, especially where a learned prior from a larger population may help the model converge given limited data. To this end, we randomly titrated the training data in the MSK dataset to 1/4, 1/16, 1/64, 1/128, 1/256, when available, and compared the performance of directly trained versus pretrained-then-finetuned SPARC models.

### Benchmarked model architectures

We compared SPARC to the following model architectures, adapted to predict the equivalent finetuning task outcomes with task-specific hyperparameter tuning using first-fold validation loss. These included those trained with only the last timepoint feature values, including MLP model ^47^, implemented using the pytorch package ^48^, XGBoost ^49^ as implemented in the scikit-learn^50^ package, as well as T-XGBoost, with input features equivalent to SPARC fed into XGBoost as features, TabPFN v3 ^51^, an in-context learning time series foundation model, as well as DuETT ^52^, where our features were averaged and binned to cater to its inputs, and Raindrop ^53^, a graphical neural network based EHR model. Significance was assessed using a paired bootstrap of the model predictions on the held-out test set (1,000 resamples), as implemented in the sci-kit learn ^50^ package. Unless otherwise stated, the principal metric reported is weighted AUROC for side - effect tasks and overall AUROC for mortality tasks for performance analyses. Weighted AUROC is the average of within-cycle AUROCs, weighted by the number of patients per cycle. This captures discrimination between patients at the same treatment stage rather than differences in baseline side-effect rates across cycles, and the weighting limits the influence of smaller later cycles. AUROC-based metrics were chosen because their standardized range allows comparison across tasks. Additional metrics, such as AUPRC, are reported in the supplementary materials.

### Downstream task clinical benchmarks

We compared SPARC to the following clinical nomograms and settings in the held-out test set of the ablation cohort where there is sufficient data required for the nomograms. We compared SPARC against each nomogram’s published coefficients, and we also refit the same features on our training and validation data, for fairness of comparison, using DeLong’s test to assess statistical significance.

- AKI: Motwani et al. ^13^ built a 4-variable additive score from age, first-course cisplatin dose, history of hypertension and serum albumin, converted to probability with the published calibration (4% baseline incidence, OR 1.49 per point). We evaluated it on each patient’s first cisplatin-containing cycle; baseline eGFR was left out, as in the original model.
- Febrile neutropenia: Aagaard et al. ^12^ built FENCE score to predict first-cycle febrile neutropenia from sex, age, cancer type, disease setting, albumin, bilirubin, eGFR, infection, number of drugs and chemotherapy classes, and we used its published regression coefficients to compute cycle-1 FN probability. As in the original study, we restricted evaluation to first cycles and excluded patients on weekly cisplatin; we also report a logistic regression refit on the same features.
- Neutropenia: Lyman et al. ^11^ built a logistic model for first-cycle severe or febrile neutropenia from age, prior chemotherapy, laboratory values (AST, alkaline phosphatase, bilirubin, GFR, WBC), cancer type, chemotherapy class and G-CSF prophylaxis. We evaluated both the published coefficients and a version refit on our training data; planned relative dose intensity was left out because it cannot be reconstructed from our records.
- Severe anemia: Dranitsaris et al. ^14^ developed a logistic model for severe chemotherapy-induced anemia (Hb <8 g/dL) in breast cancer, using age, TNM stage, baseline hemoglobin, steroid use, MCV and GFR. Due to the limited number of well labelled breast cancer samples in the test set, we applied the published odds ratios to our pan-cancer cohort, which makes it an off-population external validation with a closely matched grade-3 anemia endpoint.

### Feature importance analysis

LIME ^29^ values were used to evaluate the relative importance of each feature at each time point. For each task, we repeated the analysis with each of the ten cross-validation models on the same test set. For each fold, signed importances were averaged across test windows, and the fold-level maps were then averaged. Missingness indicators and padding were excluded from reporting. To understand the relative importance of each feature and the contribution of time series, we calculated each feature’s mean absolute importance across time points weighted by the importance of each time point.

### External validation and fine tuning

We fine tuned SPARC models at INSIGHT institutions in two ways, using the equivalent model designs and hyperparameters at MSK: site-specific and pooled. Because of limited data, each site’s patients were randomly split into three equally sized sets for training, validation and testing. In the site-specific setting, models were fine-tuned and validated on a single site’s training and validation sets. In the pooled setting, the training and validation sets of all sites were combined for fine-tuning. In both settings, performance was evaluated separately on each site’s held-out test set. The ICU mortality task was not included in external validation because ICU admission and discharge dates could not be mapped reliably. Certain tasks at certain INSIGHT institutions with low count of <20 patients were excluded in evaluation.

### G-CSF statistical analysis

SPARC predictions for the neutropenia task were generated on the MSK test set. A sample was considered G-CSF-exposed if G-CSF was administered within 2 days after the prediction cutoff date (and before its onset). Samples were stratified into tertiles of SPARC-predicted risk. The propensity of receiving G-CSF was estimated by L2-regularized logistic regression on the standardized model inputs (feature values, missingness indicators, visit times and infusion number), trained on the training set. On the test set, these scores were recalibrated against G-CSF receipt alone, without using outcomes, and then used for weighting. As the number of control samples matched with G-CSF treated samples is a hyperparameter that may change results, for the main analysis, we used inverse probability of treatment weighting (IPTW) with overlap weights (1 − e for exposed, e for unexposed infusions; e, propensity score), which down-weight extreme propensity scores. Within each tertile, we computed weighted neutropenia rates, the absolute risk reduction (ARR) and the number needed to treat (NNT = 1/ARR), with 95% CIs and two-sided p-values from a patient-level bootstrap (2,000 resamples). Effective sample sizes were computed with Kish’s formula, and positivity was assessed from propensity score overlap between exposed and unexposed infusions overall and within each tertile (**Supplementary Figure 8b**).

As a sensitivity analysis, we used propensity score matching (Supplementary Figure 8a). Each G-CSF-exposed infusion was matched to 1, 2 or 10 unexposed infusions via nearest propensity score (nearest-neighbor matching with replacement). within the same risk tertile and infusion cycle, by nearest neighbour on the logit propensity score with a caliper of 0.2 standard deviations, with replacement. NNTs, 95% confidence intervals and p-values were computed with the same patient-level bootstrap.

### Software

Data preprocessing, SPARC models, and deep learning benchmark models, unless otherwise stated, were implemented with Python 3.9.23, PyTorch Lightning 2.3.3, PyTorch 2.6.0, CUDA 12.4, NumPy 1.26.4. For additional benchmarks, we leveraged XGBoost 2.1.4 and TabPFN v3 from version 8.0.7. Models were evaluated with torchmetrics 1.5.2, SciPy 1.13.1 and statsmodels 0.14.4. Regression benchmarks for pretraining and additional metrics were implemented with scikit-learn 1.5.2.

Data preprocessing for CAIA institutions was conducted by CAIA institutional collaborators at respective institutions following the same protocol and uploaded into Rhino Federated Computing Platform, where SPARC performance metrics were acquired via zero-shot inference. Upon data requests, INSIGHT CRN’s provided OMOP-format data was processed into formats aligned with CAIA datasets, for further analyses.

## Data Availability Statement

SPARC model weights and codes are available under the Creative Commons BY-NC-ND 4.0 licence and will be accessible on HuggingFace upon formal publication. Data from the EHRSHOT and MIMIC IV-hosp cohorts need to be requested at https://physionet.org and following the corresponding instructions, and can then be pre-processed to form SPARC input format leveraging codes in our github repository. Questions regarding data requests from INSIGHT-CRN should be directed to https://insightcrn.org/data-overview/access-our-data/ while CAIA data and collaborations, including MSK data, should be addressed to Brian Bot.

## Code Availability

Code can be found at https://github.com/tommyfuu/SPARC. Model weights for the pretrained and fine-tuned SPARC-P models are hosted at https://huggingface.co/tommyfuu/SPARC.

## Acknowledgement

We acknowledge the contributions of the US Cancer AI Alliance (CAIA) and the INSIGHT Clinical Research Network, as well as their participating institutions, for their contribution to the project. We thank Kristen Tomick for her contributions to the organization of CAIA. This work was supported by a Walther Foundation ASCO Conquer Cancer Career Development Award, the Society of MSK, and The National Cancer Institute (K08CA286842, P30 CA008748).

## Author contribution

C.F., Q.M., and J.J. designed and performed experiments, analyzed data, and wrote the manuscript. J.Y., R.S., M.W., A.A., S.P.S., K.L.K., B.M.B., and N.S. contributed to data processing and analysis. M.W., M.P., S.P.S., A.B., K.L.K, B.M.B, and N.S. supported or provided the access to institutional datasets via the CAIA consortium. Q.M., N.S., and J.J. conceived the study, conceptualized the project and supervised the research. All authors edited the manuscript, reviewed and approved the paper.

## CAIA Consortium

**Dana-Farber Cancer Institute, Boston, MA, USA** — Jason Johnson, Kevin Haigis, Amit Kulkarni, Ethan Cerami, John Methot, Michael Rosenthal, Sherry Lee, Susan Barry, Thejas Bharadwaj, Sarah Paulino, Stephen Dollinger, Eliezer Van Allen, Xuelu (Jeff) Liu, Yantong Cui & Mark Apinis

**Fred Hutchinson Cancer Center, Seattle, WA, USA** — Jeff Leek, Bruce Clurman, Brian M. Bot, Amy Paguirigan, Monica Gerber, Anna Oldre, Josh Southam & Megan Stachura

**Johns Hopkins University, Baltimore, MD, USA** — Srinivasan Yegnasubramanian, Alexis Battle, Alex Baras, Edward Hunter, Jeff Weaver, Richard Mendola, Benjamin Martin, James Weatherell, Taxiarchis Botsis, Phil Taft, Sudip Baniya, Jorge Marquez & Resford Rouzer

**Memorial Sloan Kettering Cancer Center, New York, NY, USA** — Sohrab Shah, Stephen Cleaver, Arfath Pasha, Julian Schwartz, Sarah Manglicmot, Shantha Bethusamy, Tatyana Sandler, Thomas Kim, Michele Waters, Alexander Hollingsworth, Anca Comanescu, Avijit Chatterjee, Javier Villarreal Almanza, Kiran Enumala, Walee Ahmed, Shakeeb Akhter, Lena Aljehane & Nikolaus Schultz

