## Supplemental materials and figures for "Longitudinal AI Models Predict Cancer Adverse Events"

Table of content

1. Supplemental Descriptions and Methods
    1.1 Detailed descriptions of ablation studies
    1.2 Cancer type assignment
2. Supplemental Figures.

### 1. Supplemental Descriptions and Methods

#### *1.1 Detailed descriptions of ablation studies*

A series of ablation studies were conducted to optimize model designs using the 57,510-patient ablation cohort, already briefly described in the Results section with results visualized in Supplemental Figure 4. Here we describe in more detail.

- Parsimony experiment: we first evaluated whether the most routine (parsimonious) lab and vital features are sufficient in predicting our outcomes accurately, or the additional modalities were crucial. We compared directly trained (no pretraining) SPARC models trained with the parsimonious features as implemented in our final model, including one-hot encoded sex, 5-year binned age, and the parsimonious lab and vital signs features, as well as SPARC models of equivalent architecture with all features. To retain the modular designs, in addition to the parsimonious models' included feature modules, the all-feature models additional include the following features fed in in the form of modules: tumour markers (Alphafetoprotein, CEA, PSA, CA-125, CA 15-3, CA 19-9, Beta-2 Microglobulin, TSH), immature and waste blood cells (Blast, Promyelo, Myelo, Meta, Immature Granulocyte, Luc, Variant Lymph, Megakaryocyte Fragment, Nucleated RBC, Conjugated Bili), mutation (genomic  features), clinical (Yost index, ECOG, genetic ancestry, smoking status, primary cancer type).
- Modular experiment: we compared directly trained SPARC models with the modularized feed-forward networks, as implemented in our final SPARC model, to ones without, trained and evaluated on equivalent data.
- Medication-as-a-feature experiment: for each side effect prediction task, we compared our SPARC models which did not explicitly encode medications as input features, as consistent with our final design, to a version of the models with one-hot encoded the medication(s) a patient is taking at cut-off given the entire list of medications for that task, as defined in *Description of prediction tasks* in *Methods* included as additional features, trained and evaluated on equivalent data.
- Medication-specific experiment: for each side effect prediction task, we compared SPARC models trained with patients receiving one or more of all medications defined in *Description of prediction tasks* in *Methods* for the task, as consistent with our final design,  to SPARC models trained with patients treated with one medication at a time, and evaluated on that medication for the fairness of comparison.
- Loss function optimization experiment: for each task, we compared SPARC models of equivalent architecture, but trained with binary cross entropy loss, Cox loss, or a hybrid of both. For the latter two, we consider the number of infusions from current time to event or the lack thereof as the time in the Cox analysis. The hybrid objective combined the Cox negative partial log-likelihood (Efron ties) with a binary cross-entropy term on next-infusion event occurrence, $L = L\_Cox + 20 \cdot L\_BCE$, where the BCE target was 1 if the adverse event occurred at the immediately following infusion and 0 otherwise.

Given the results of these experiments as discussed in the **Results** section, upon pretraining, the pretrained SPARC weights are then used as weight initializations to be finetuned for downstream tasks based on the parsimonious, modular, Cox or hybrid setups without including medications as features.

*1.5 Cancer type assignment*

Patients were mapped to one (or multiple) of the following primary cancer types given their earliest diagnosed cancer condition code (ICD billing codes for INSIGHT, MIMIC IV-hosp, and EHRSHOT, and OMOP condition codes for CAIA institutions): Appendix Cancer, Bladder Cancer, Bone Cancer, Breast Cancer, Cervical Cancer, Colorectal Cancer, Endometrial Cancer, Esophagogastric Cancer, GIST, Germ Cell Tumor, Glioma, Head and Neck Cancer, Hepatobiliary Cancer, Leukemia, Lymphoma, Mature T and NK Neoplasms, Mesothelioma, Myelodysplastic Syndromes, Neuroendocrine Tumor, Non-Small Cell Lung Cancer, Ovarian Cancer, Pancreatic Cancer, Prostate Cancer, Renal Cell Carcinoma, Sarcoma, Skin Cancer, Small Cell Lung Cancer, Thyroid Cancer, Unknown Primary, Uterine Sarcoma. Codes for in-situ lesions and metastatic cancers are excluded, so patients with only such codes will have missing cancer type mapping, despite their inclusion in the cohort.

#### 2. Supplemental Figures

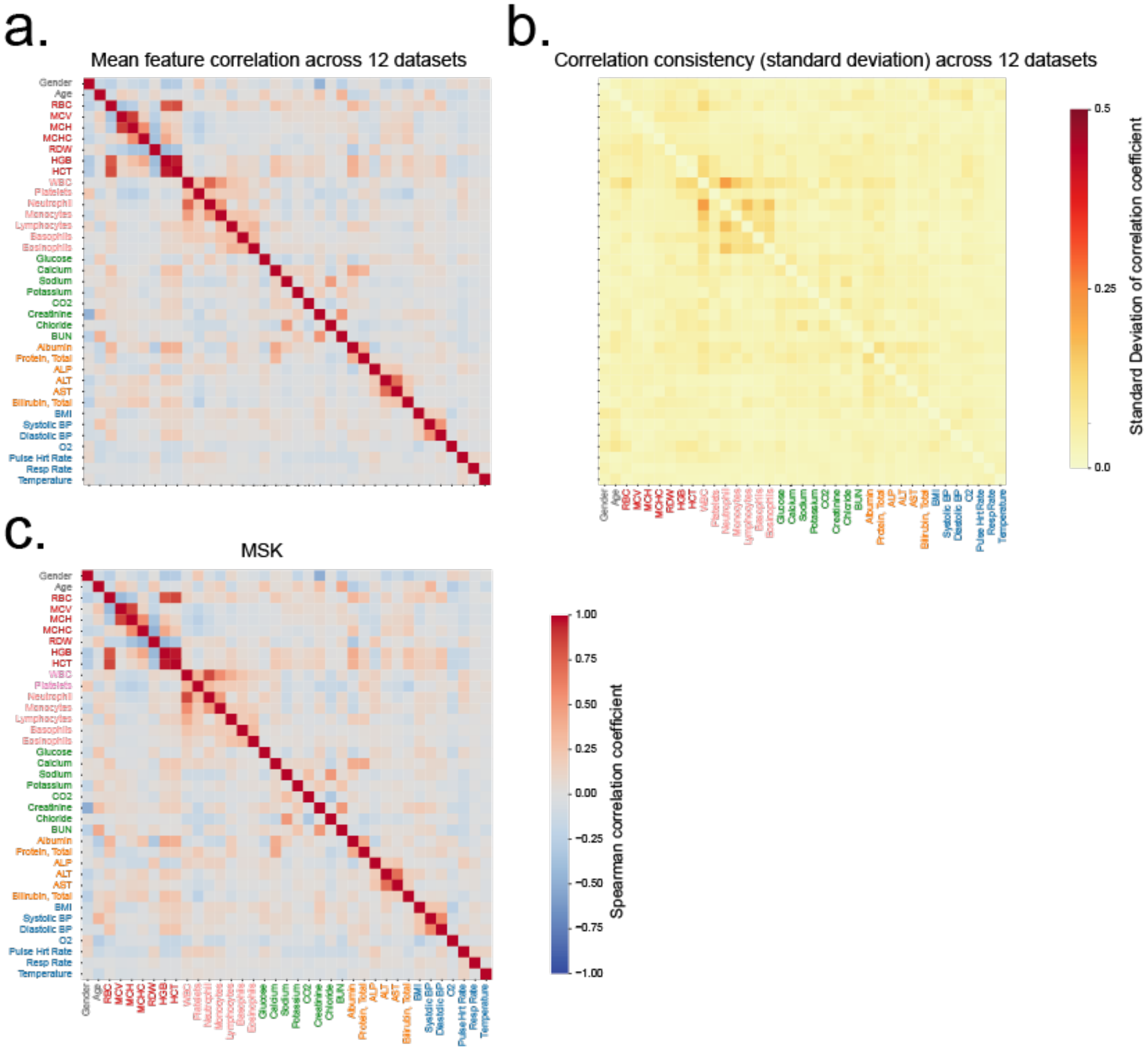

**Supplemental Figure 1. Module-wise feature correlation.**
We computed and observed the feature correlation across features using first lab value per feature per patient within the MSK cohort (c) as well as across 12 independently collected cohorts with cancer (See Methods for methodology and Supplemental Table 1 for demographics).

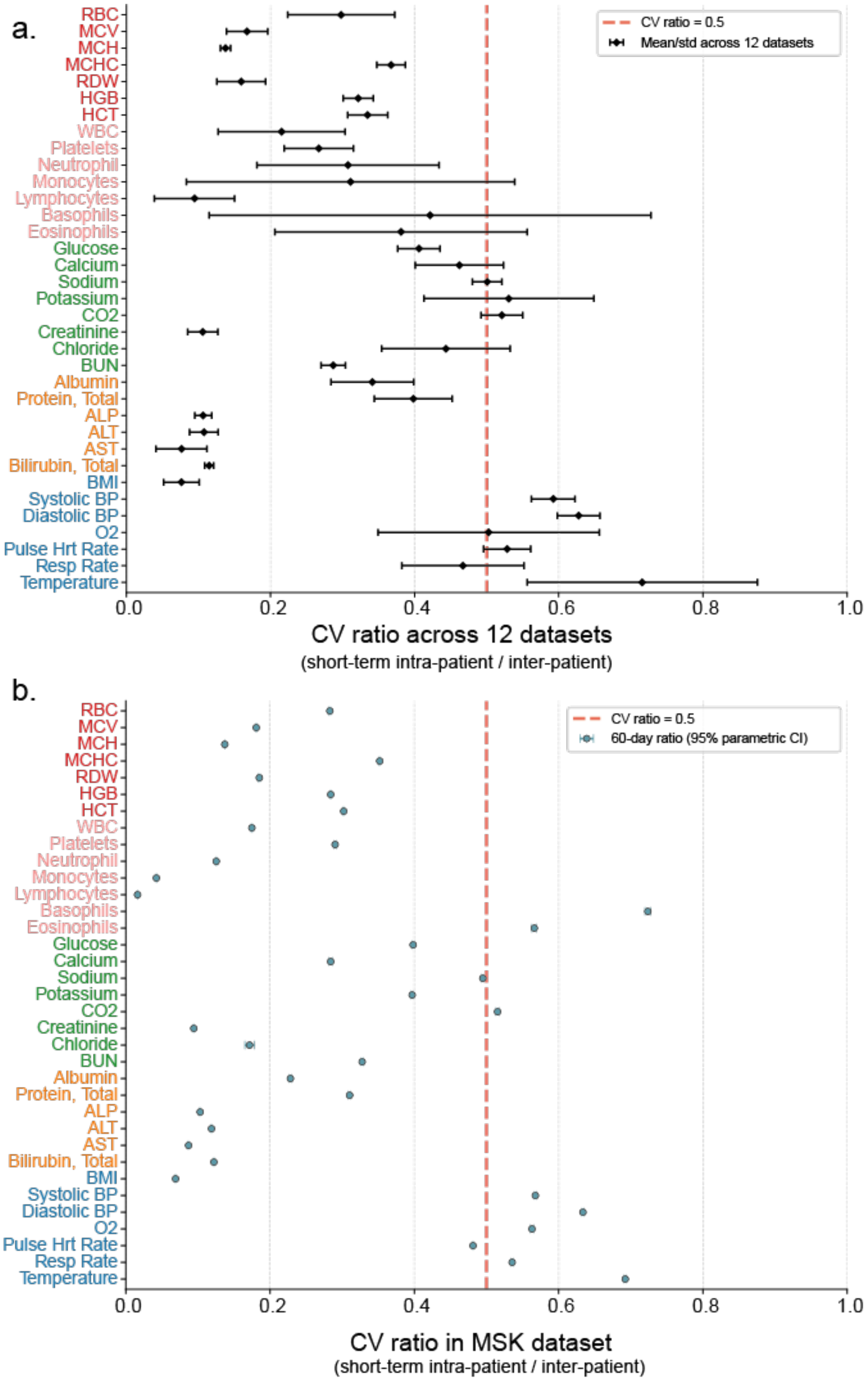

**Supplemental Figure 2. CV ratio within MSK and across validation datasets.**
We computed and observed the ratio between CV within and across patients across features within the MSK cohort (c) as well as across 12 independently collected cohorts with cancer (See Methods for methodology and Supplemental Table 1 for demographics).

a.

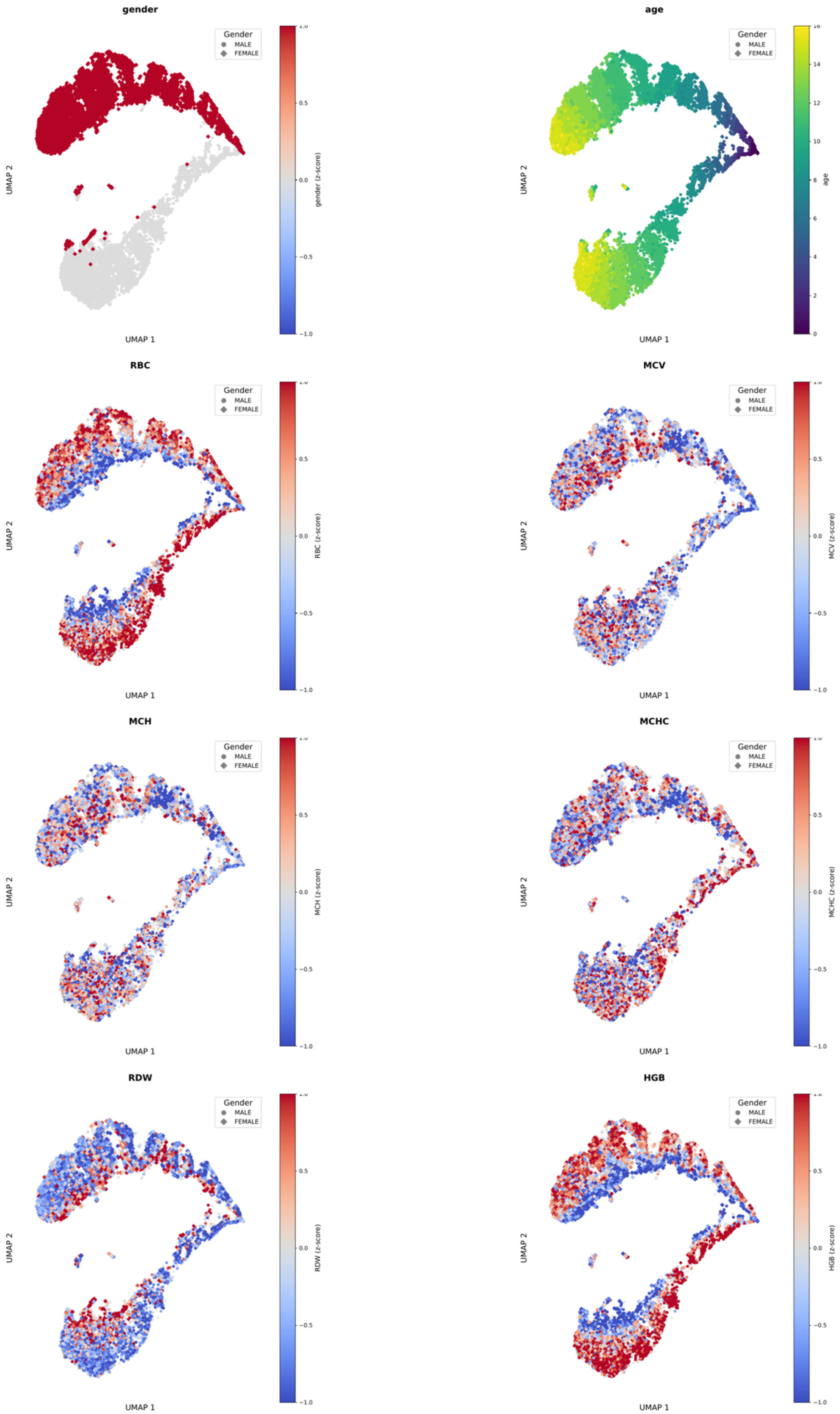

gender
age
RBC
MCV
MCH
MCHC
RDW
HGB
Gender
MALE
FEMALE
UMAP 1
UMAP 2
gender (z-score)
age
RBC (z-score)
MCV (z-score)
MCH (z-score)
MCHC (z-score)
RDW (z-score)
HGB (z-score)

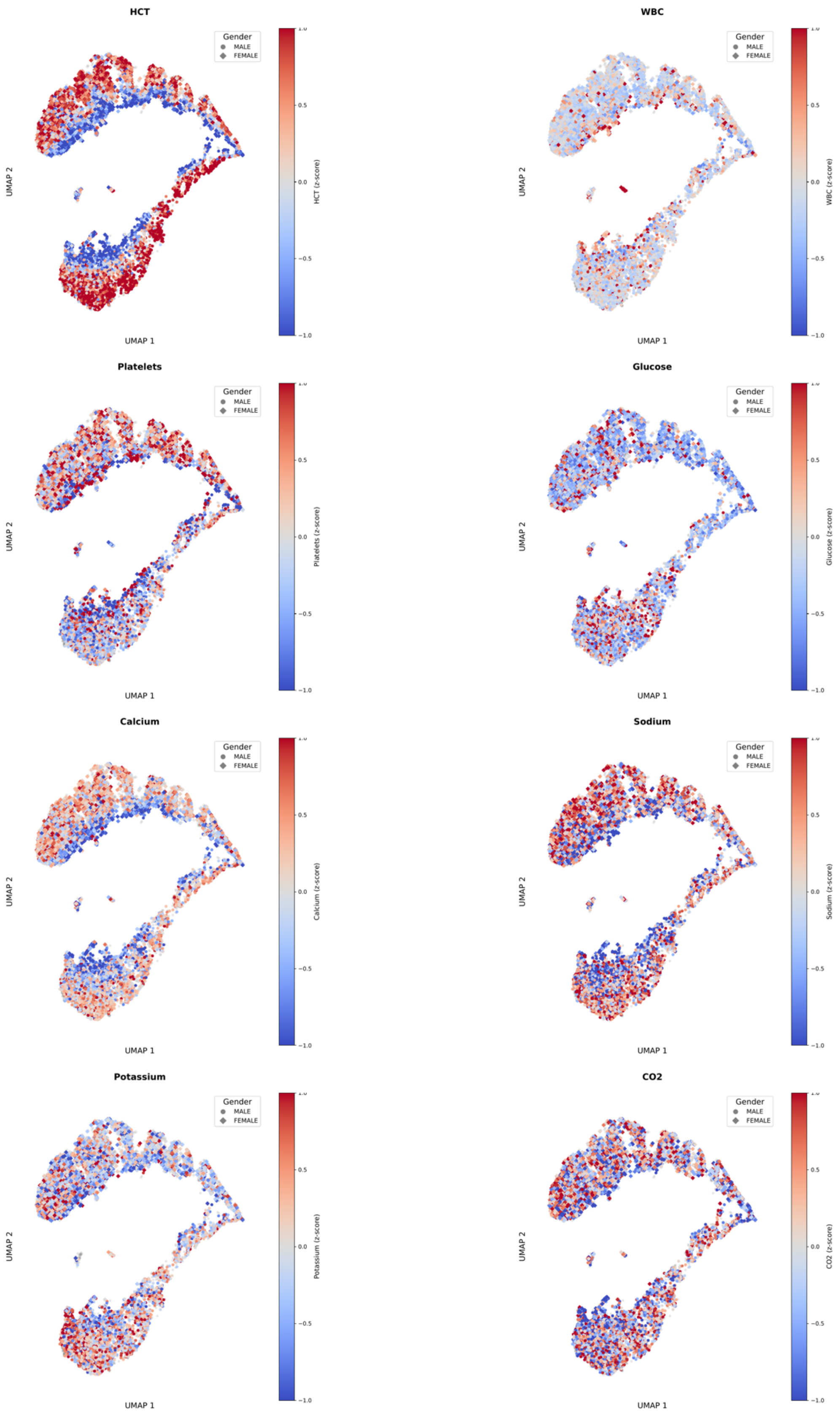

HCT
Gender
MALE
FEMALE
UMAP 2
UMAP 1
HCT (z-score)
WBC
WBC (z-score)
Platelets
Platelets (z-score)
Glucose
Glucose (z-score)
Calcium
Calcium (z-score)
Sodium
Sodium (z-score)
Potassium
Potassium (z-score)
CO2
CO2 (z-score)
1.0
0.5
0.0
−0.5
−1.0

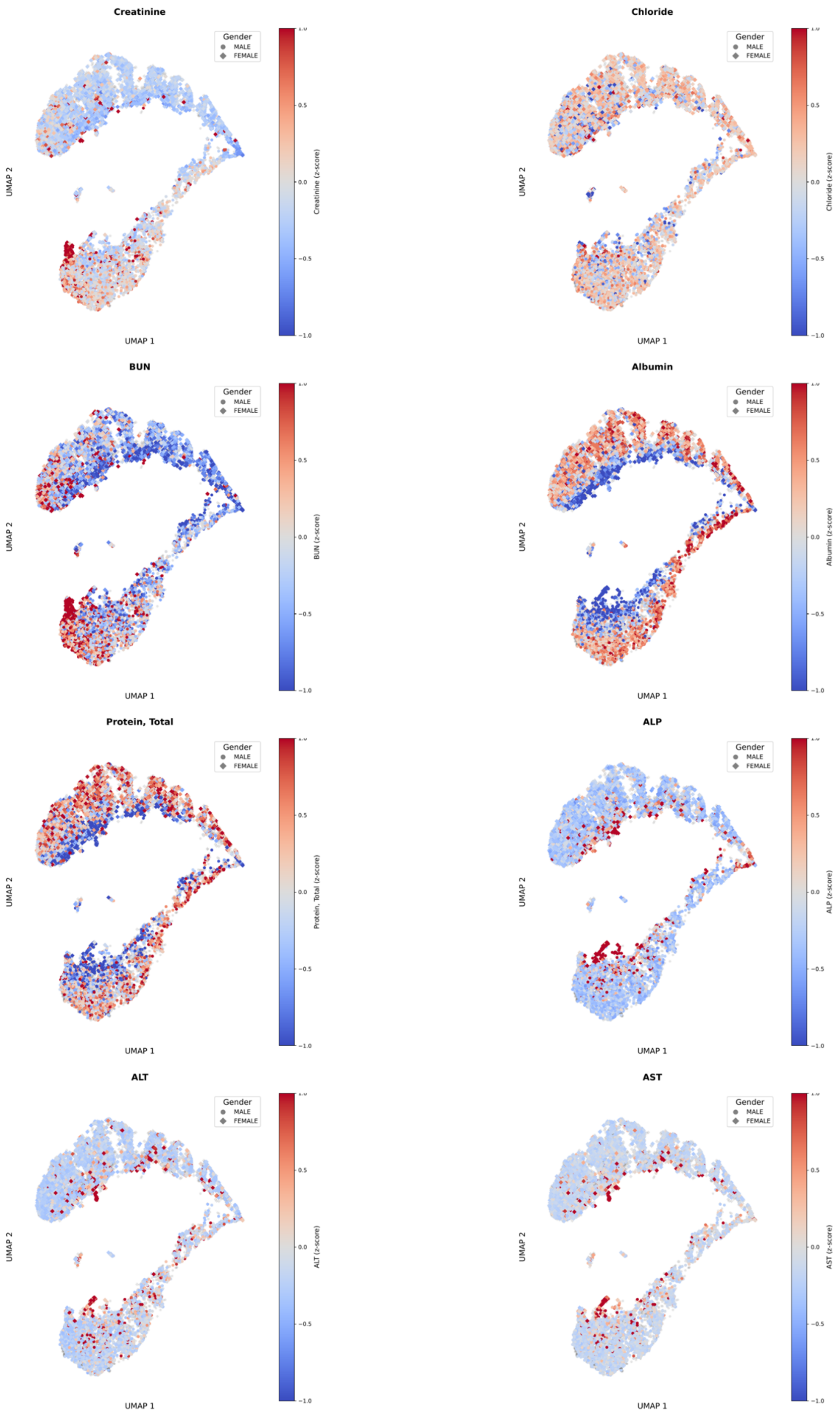

Creatinine
Gender
MALE
FEMALE
UMAP 1
UMAP 2
Creatinine (z-score)
Chloride
Chloride (z-score)
BUN
BUN (z-score)
Albumin
Albumin (z-score)
Protein, Total
Protein, Total (z-score)
ALP
ALP (z-score)
ALT
ALT (z-score)
AST
AST (z-score)
1.0
0.5
0.0
−0.5
−1.0

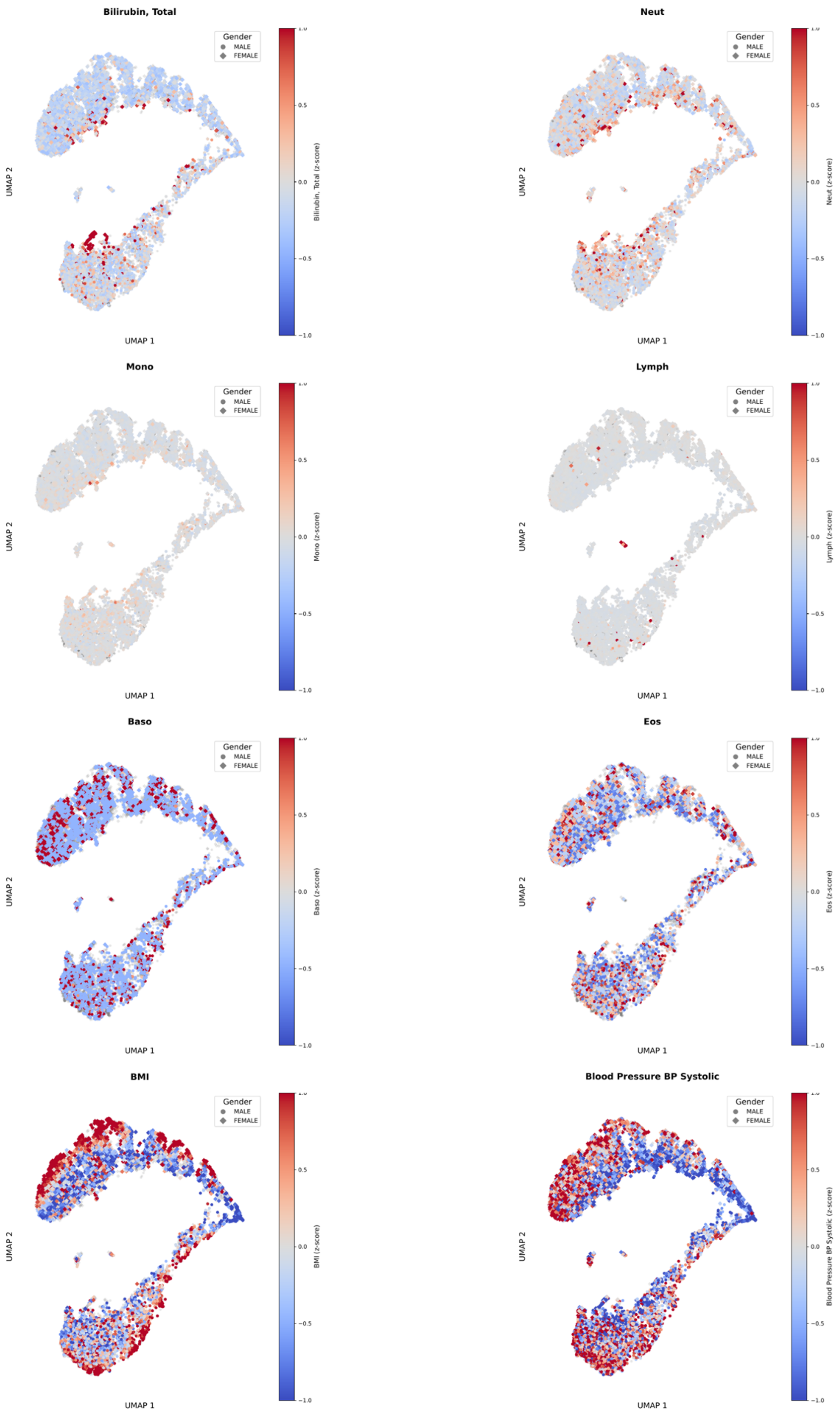

Bilirubin, Total
Gender
MALE
FEMALE
UMAP 2
UMAP 1
Bilirubin, Total (z-score)
1.0
0.5
0.0
−0.5
−1.0
Neut
Neut (z-score)
Mono
Mono (z-score)
Lymph
Lymph (z-score)
Baso
Baso (z-score)
Eos
Eos (z-score)
BMI
BMI (z-score)
Blood Pressure BP Systolic
Blood Pressure BP Systolic (z-score)

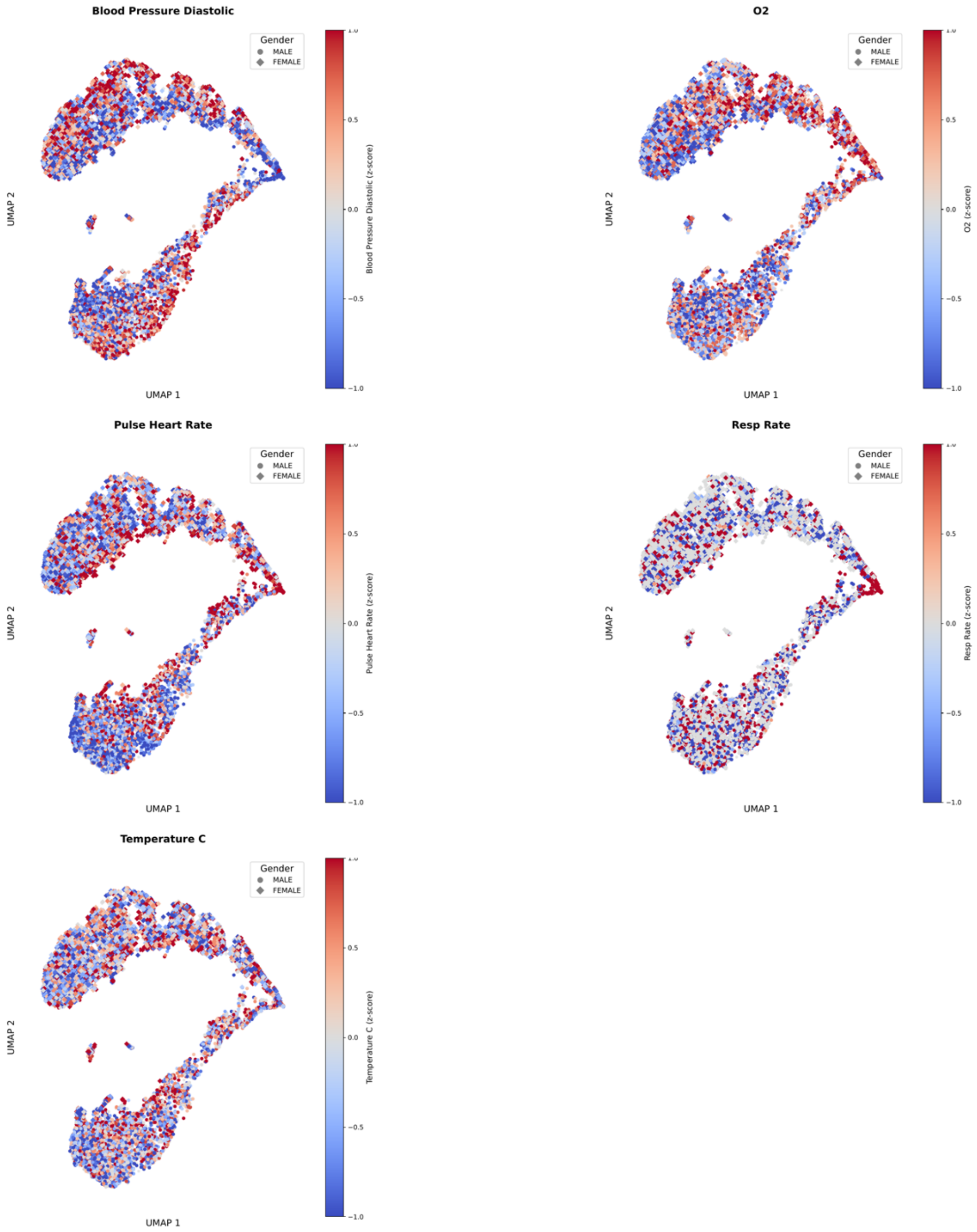
Blood Pressure Diastolic
Gender
MALE
FEMALE
UMAP 2
UMAP 1
Blood Pressure Diastolic (z-score)
1.0
0.5
0.0
−0.5
−1.0
O2
Gender
MALE
FEMALE
UMAP 2
UMAP 1
O2 (z-score)
Pulse Heart Rate
Gender
MALE
FEMALE
UMAP 2
UMAP 1
Pulse Heart Rate (z-score)
Resp Rate
Gender
MALE
FEMALE
UMAP 2
UMAP 1
Resp Rate (z-score)
Temperature C
Gender
MALE
FEMALE
UMAP 2
UMAP 1
Temperature C (z-score)

b.

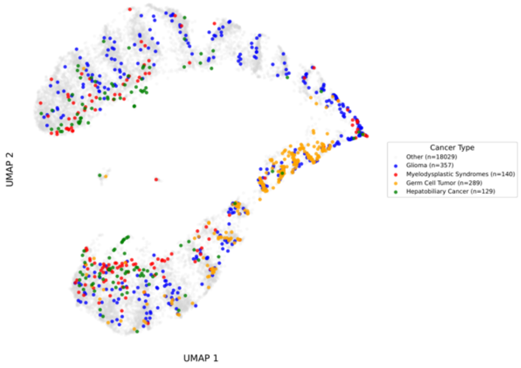

**Supplemental Figure 3. Pretrained patient-specific embeddings.**

UMAP visualizations of the pretrained embeddings of their randomly selected pretraining windows for patients in the MSK-CAIA held-out test set are visualized, and coloured with (a) z-score normalized lab values at the predicted timepoint, and (b) cancer types. In (a), age was visualized in 5-year age groups for data anonymization purposes and thus the scale between 0 and 20.

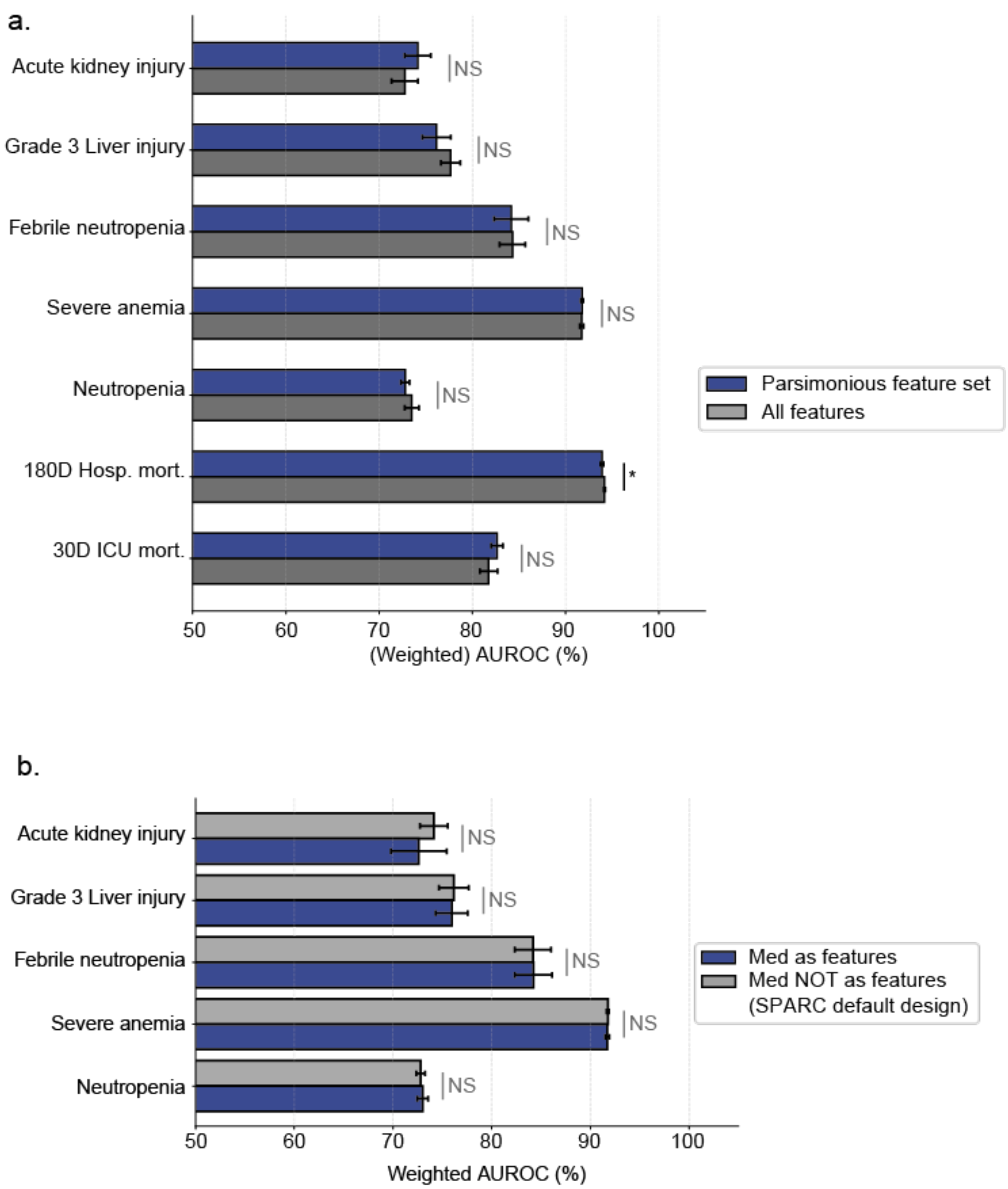
a.
Acute kidney injury
Grade 3 Liver injury
Febrile neutropenia
Severe anemia
Neutropenia
180D Hosp. mort.
30D ICU mort.
NS
*
50
60
70
80
90
100
(Weighted) AUROC (%)
Parsimonious feature set
All features
b.
Acute kidney injury
Grade 3 Liver injury
Febrile neutropenia
Severe anemia
Neutropenia
NS
50
60
70
80
90
100
Weighted AUROC (%)
Med as features
Med NOT as features
(SPARC default design)

c.

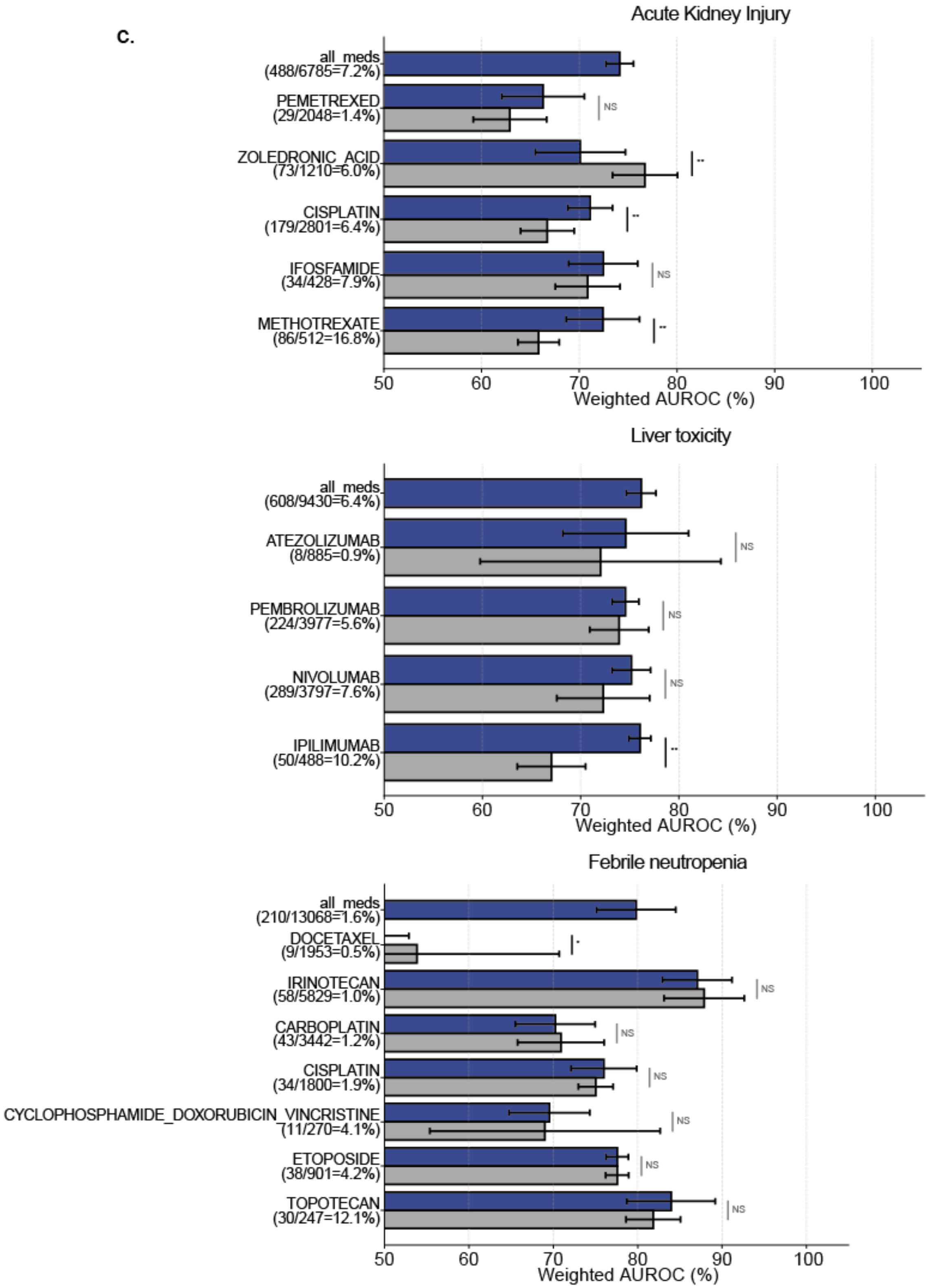

Acute Kidney Injury
all_meds (488/6785=7.2%)
PEMETREXED (29/2048=1.4%)
ZOLEDRONIC_ACID (73/1210=6.0%)
CISPLATIN (179/2801=6.4%)
IFOSFAMIDE (34/428=7.9%)
METHOTREXATE (86/512=16.8%)
NS
**
**
NS
**
50
60
70
80
90
100
Weighted AUROC (%)
Liver toxicity
all_meds (608/9430=6.4%)
ATEZOLIZUMAB (8/885=0.9%)
PEMBROLIZUMAB (224/3977=5.6%)
NIVOLUMAB (289/3797=7.6%)
IPILIMUMAB (50/488=10.2%)
NS
NS
NS
**
Weighted AUROC (%)
Febrile neutropenia
all_meds (210/13068=1.6%)
DOCETAXEL (9/1953=0.5%)
IRINOTECAN (58/5829=1.0%)
CARBOPLATIN (43/3442=1.2%)
CISPLATIN (34/1800=1.9%)
CYCLOPHOSPHAMIDE_DOXORUBICIN_VINCRISTINE (11/270=4.1%)
ETOPOSIDE (38/901=4.2%)
TOPOTECAN (30/247=12.1%)
*
NS
NS
NS
NS
NS
NS
Weighted AUROC (%)

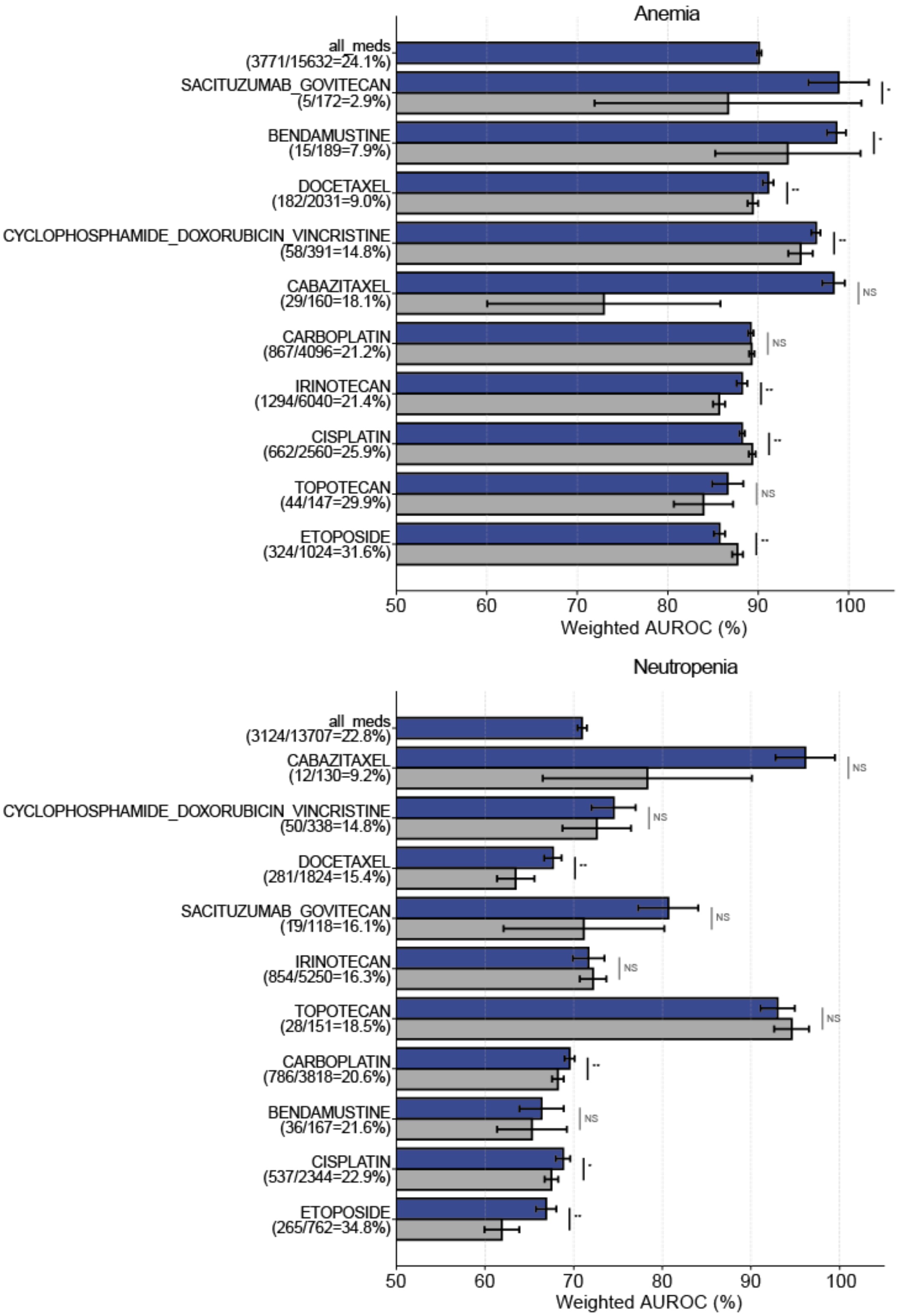
Anemia
all_meds
(3771/15632=24.1%)
SACITUZUMAB_GOVITECAN
(5/172=2.9%)
BENDAMUSTINE
(15/189=7.9%)
DOCETAXEL
(182/2031=9.0%)
CYCLOPHOSPHAMIDE_DOXORUBICIN_VINCRISTINE
(58/391=14.8%)
CABAZITAXEL
(29/160=18.1%)
CARBOPLATIN
(867/4096=21.2%)
IRINOTECAN
(1294/6040=21.4%)
CISPLATIN
(662/2560=25.9%)
TOPOTECAN
(44/147=29.9%)
ETOPOSIDE
(324/1024=31.6%)
NS
50
60
70
80
90
100
Weighted AUROC (%)
Neutropenia
all_meds
(3124/13707=22.8%)
CABAZITAXEL
(12/130=9.2%)
CYCLOPHOSPHAMIDE_DOXORUBICIN_VINCRISTINE
(50/338=14.8%)
DOCETAXEL
(281/1824=15.4%)
SACITUZUMAB_GOVITECAN
(19/118=16.1%)
IRINOTECAN
(854/5250=16.3%)
TOPOTECAN
(28/151=18.5%)
CARBOPLATIN
(786/3818=20.6%)
BENDAMUSTINE
(36/167=21.6%)
CISPLATIN
(537/2344=22.9%)
ETOPOSIDE
(265/762=34.8%)
NS
50
60
70
80
90
100
Weighted AUROC (%)

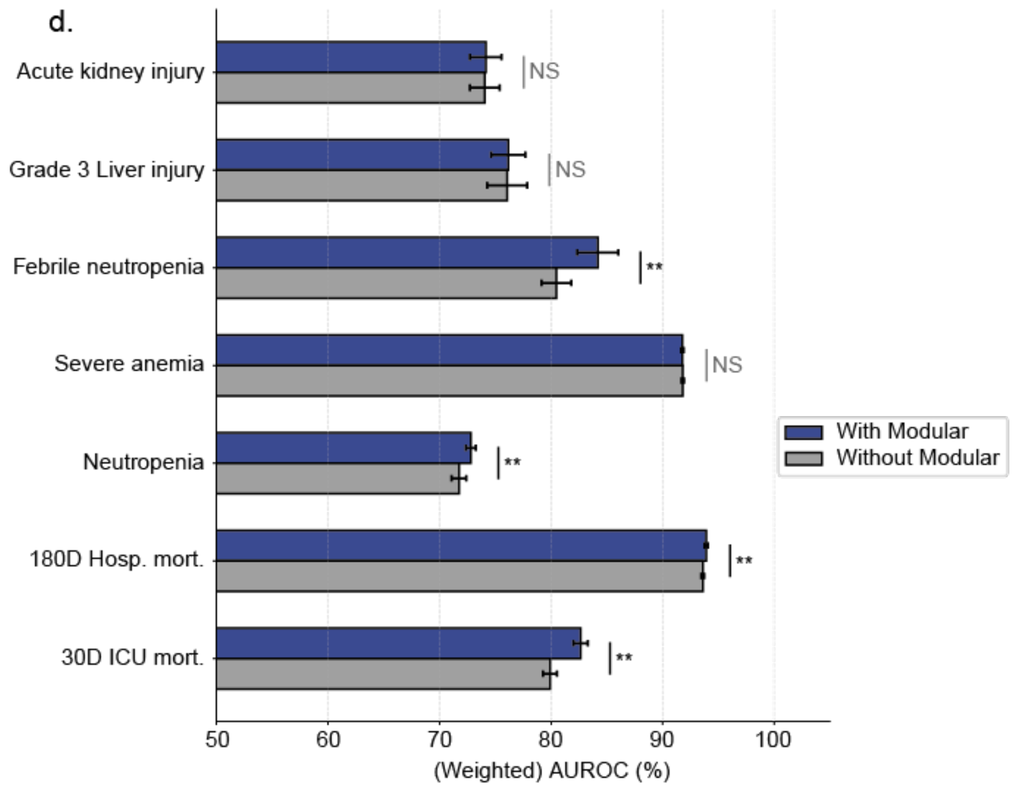
d.
Acute kidney injury
Grade 3 Liver injury
Febrile neutropenia
Severe anemia
Neutropenia
180D Hosp. mort.
30D ICU mort.
NS
NS
**
NS
**
**
**
With Modular
Without Modular
50
60
70
80
90
100
(Weighted) AUROC (%)

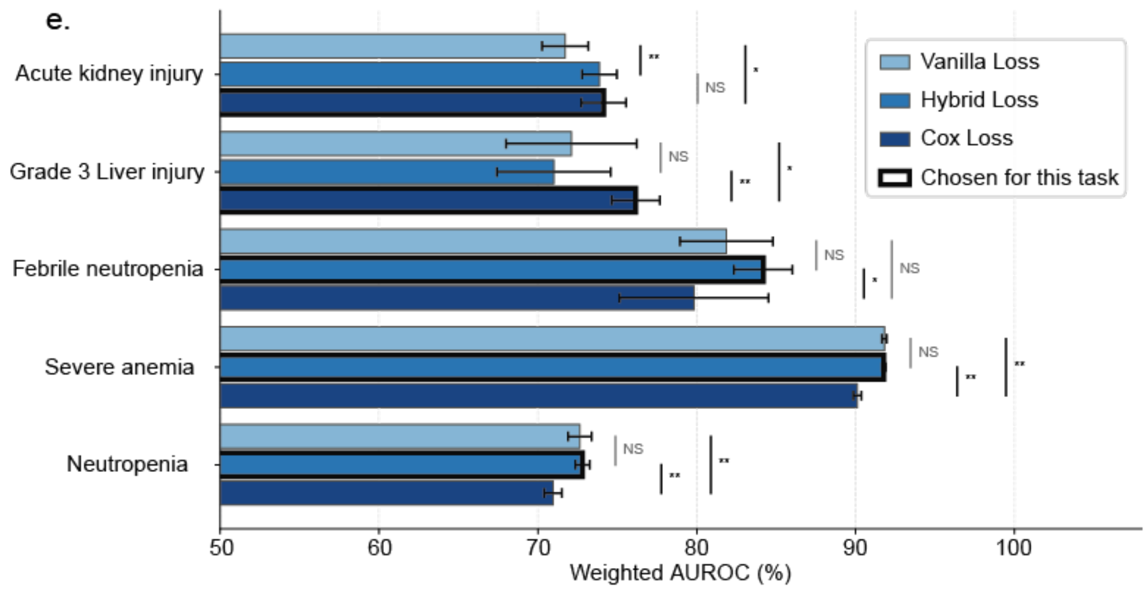
e.
Acute kidney injury
Grade 3 Liver injury
Febrile neutropenia
Severe anemia
Neutropenia
Vanilla Loss
Hybrid Loss
Cox Loss
Chosen for this task
50
60
70
80
90
100
Weighted AUROC (%)

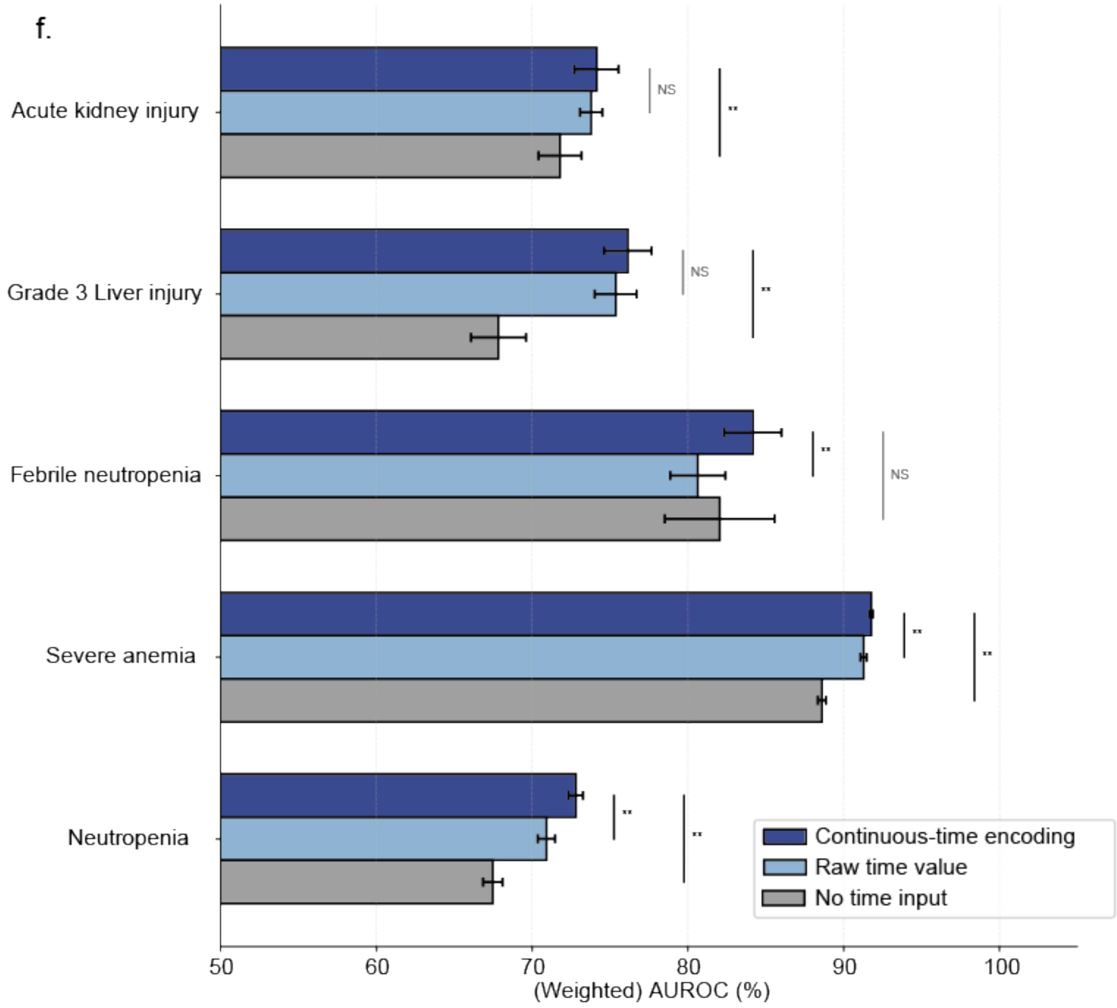

**Supplemental Figure 4. Ablation studies for SPARC design choice decisions.**
We compared our SPARC models in a directly supervised setting to equivalent models with one design component removed or altered at a time: **(a)** parsimonious feature set (in finalized SPARC) versus all features (see Methods for definition); **(b)** included medications not as features (in finalized SPARC) versus included medications one-hot encoded as features (see Methods for definition); **(c)** pooled training across medications (in finalized SPARC) versus medication-specific model; **(d)** with module-specific embeddings (in finalized SPARC) versus without; **(e)** using supervised loss with a Cox loss component (in finalized SPARC) versus without; and **(f)** with continuous-time encoding (in finalized SPARC) versus directly inputting time as a feature without embedding and not using time as a feature (see Methods for definition).

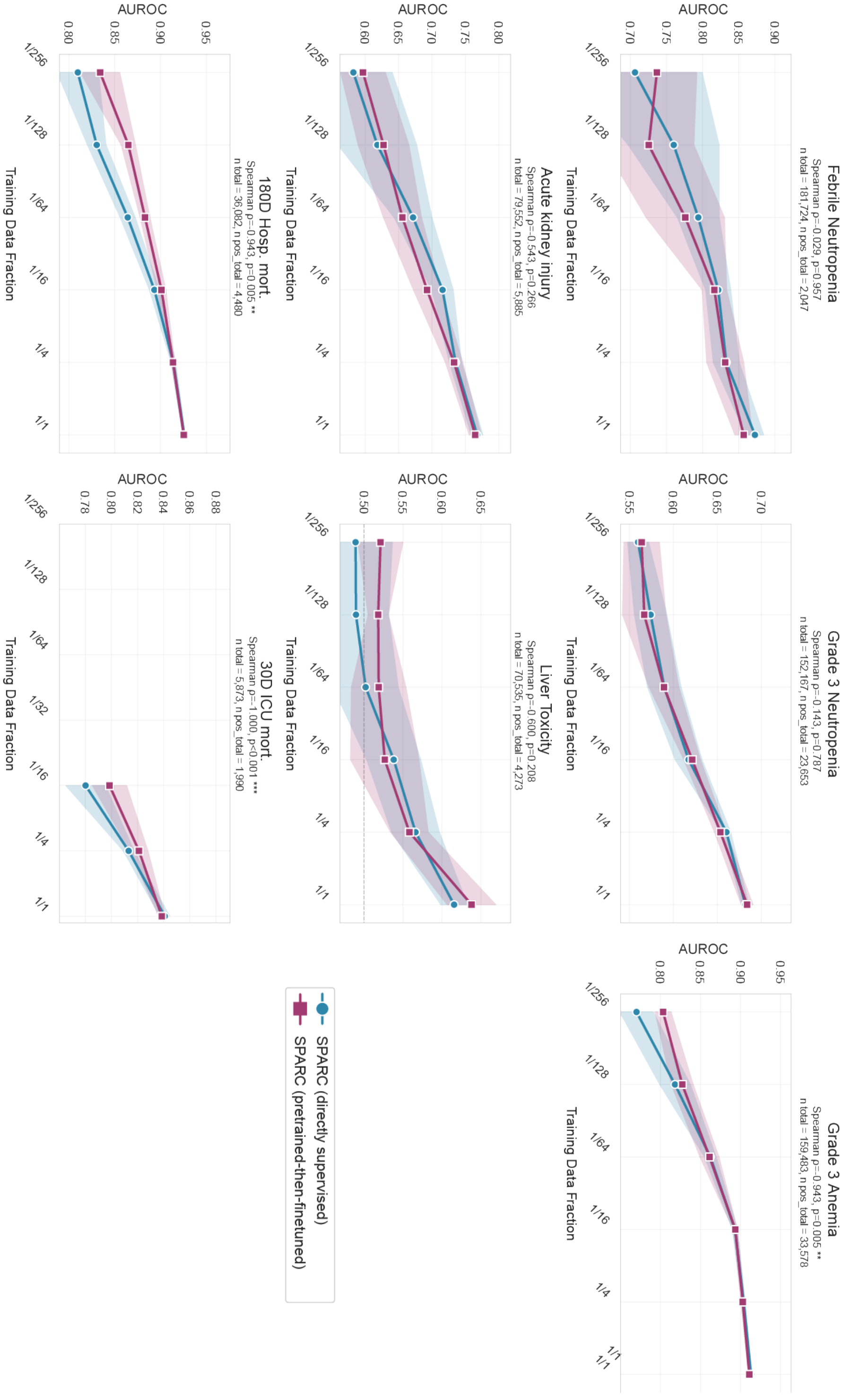

**Supplemental Figure 5. Performance of pretrained-then-finetuned and directly supervised SPARC models in different fractions of training data.** See Methods for Methodology.

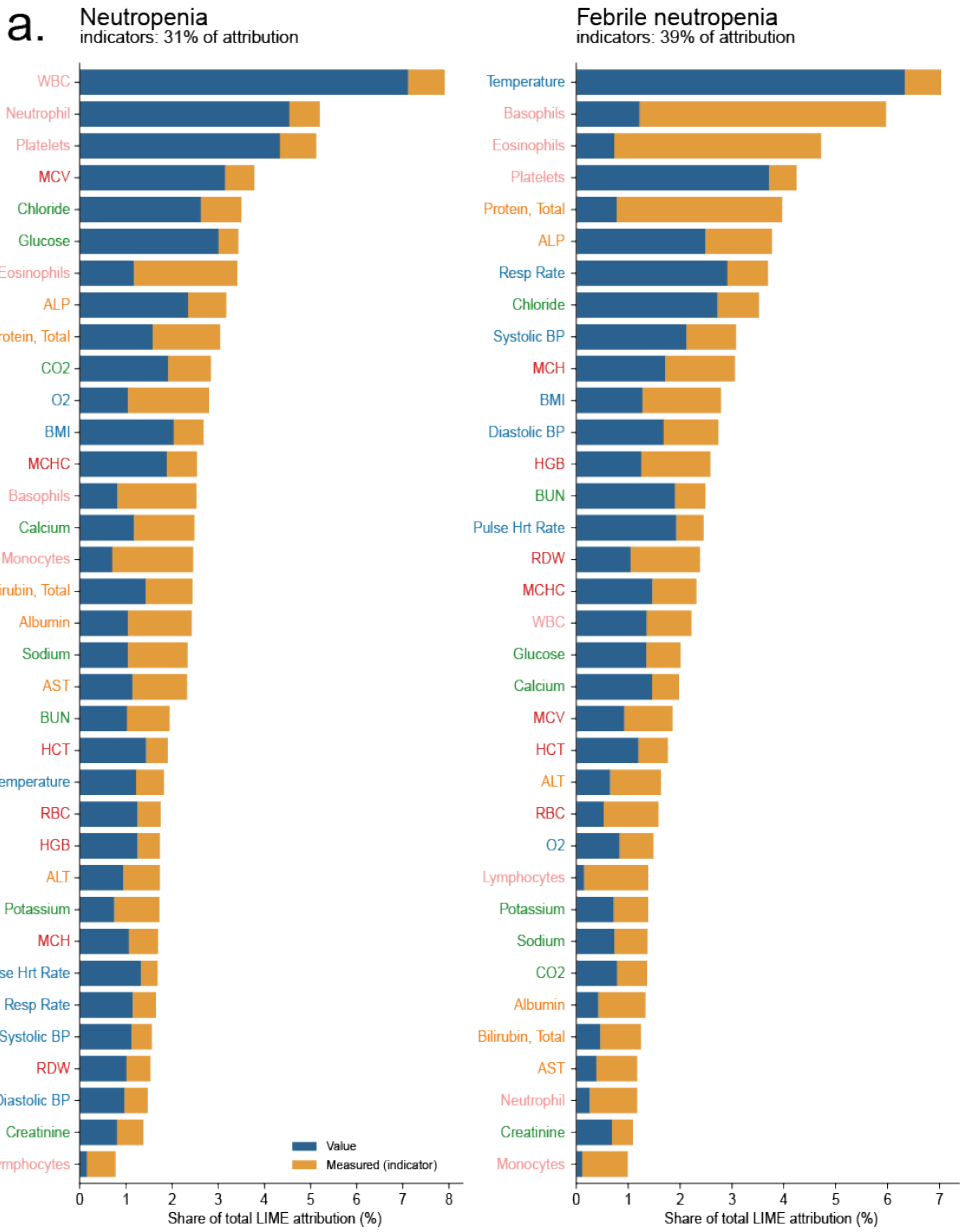

a.
Neutropenia
indicators: 31% of attribution
WBC
Neutrophil
Platelets
MCV
Chloride
Glucose
Eosinophils
ALP
Protein, Total
CO2
O2
BMI
MCHC
Basophils
Calcium
Monocytes
Bilirubin, Total
Albumin
Sodium
AST
BUN
HCT
Temperature
RBC
HGB
ALT
Potassium
MCH
Pulse Hrt Rate
Resp Rate
Systolic BP
RDW
Diastolic BP
Creatinine
Lymphocytes
Value
Measured (indicator)
0 1 2 3 4 5 6 7 8
Share of total LIME attribution (%)
Febrile neutropenia
indicators: 39% of attribution
Temperature
Basophils
Eosinophils
Platelets
Protein, Total
ALP
Resp Rate
Chloride
Systolic BP
MCH
BMI
Diastolic BP
HGB
BUN
Pulse Hrt Rate
RDW
MCHC
WBC
Glucose
Calcium
MCV
HCT
ALT
RBC
O2
Lymphocytes
Potassium
Sodium
CO2
Albumin
Bilirubin, Total
AST
Neutrophil
Creatinine
Monocytes
0 1 2 3 4 5 6 7
Share of total LIME attribution (%)

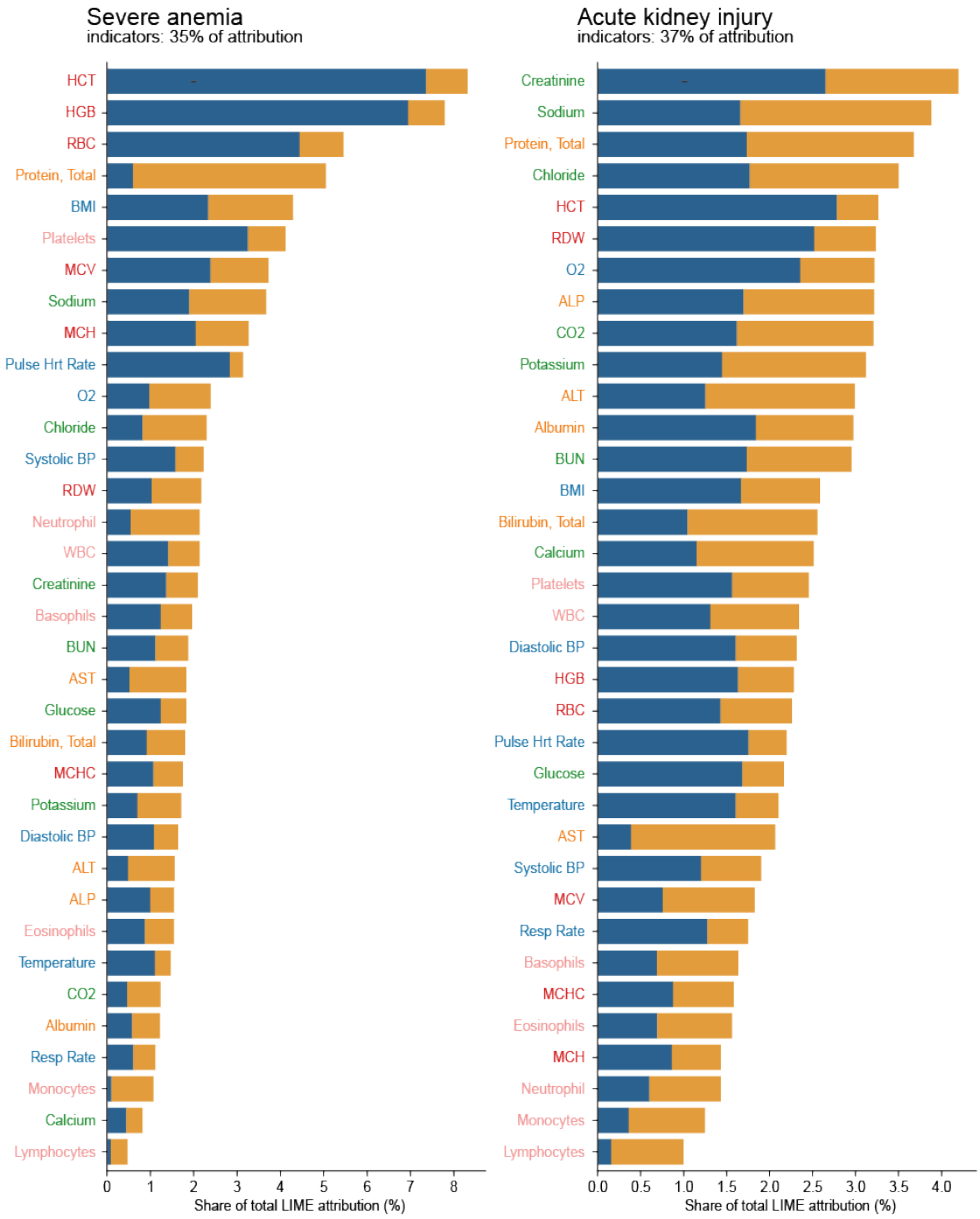

Severe anemia
indicators: 35% of attribution
HCT
HGB
RBC
Protein, Total
BMI
Platelets
MCV
Sodium
MCH
Pulse Hrt Rate
O2
Chloride
Systolic BP
RDW
Neutrophil
WBC
Creatinine
Basophils
BUN
AST
Glucose
Bilirubin, Total
MCHC
Potassium
Diastolic BP
ALT
ALP
Eosinophils
Temperature
CO2
Albumin
Resp Rate
Monocytes
Calcium
Lymphocytes
0
1
2
3
4
5
6
7
8
Share of total LIME attribution (%)
Acute kidney injury
indicators: 37% of attribution
Creatinine
Sodium
Protein, Total
Chloride
HCT
RDW
O2
ALP
CO2
Potassium
ALT
Albumin
BUN
BMI
Bilirubin, Total
Calcium
Platelets
WBC
Diastolic BP
HGB
RBC
Pulse Hrt Rate
Glucose
Temperature
AST
Systolic BP
MCV
Resp Rate
Basophils
MCHC
Eosinophils
MCH
Neutrophil
Monocytes
Lymphocytes
0.0
0.5
1.0
1.5
2.0
2.5
3.0
3.5
4.0
Share of total LIME attribution (%)

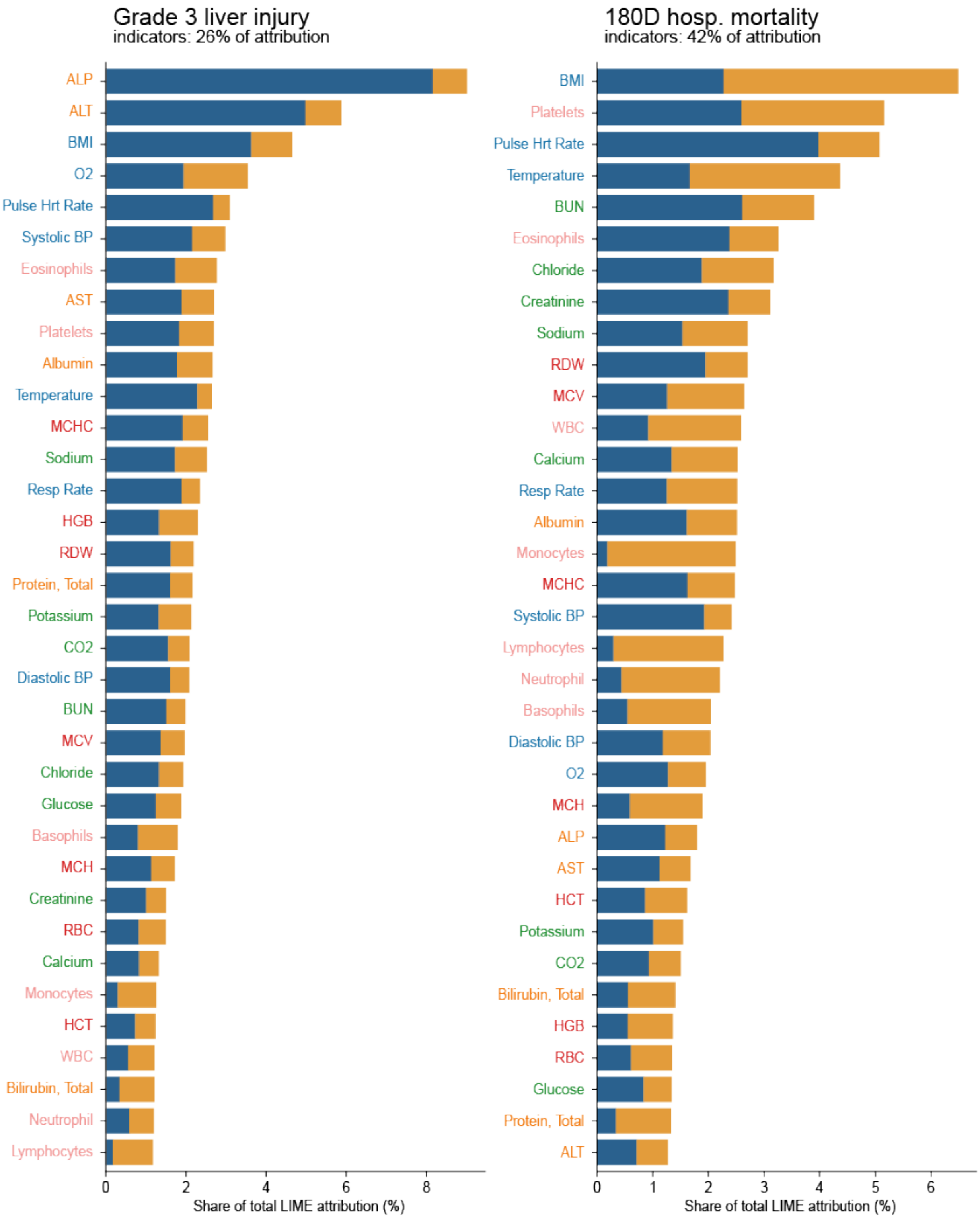

Grade 3 liver injury
indicators: 26% of attribution
ALP
ALT
BMI
O2
Pulse Hrt Rate
Systolic BP
Eosinophils
AST
Platelets
Albumin
Temperature
MCHC
Sodium
Resp Rate
HGB
RDW
Protein, Total
Potassium
CO2
Diastolic BP
BUN
MCV
Chloride
Glucose
Basophils
MCH
Creatinine
RBC
Calcium
Monocytes
HCT
WBC
Bilirubin, Total
Neutrophil
Lymphocytes
0
2
4
6
8
Share of total LIME attribution (%)
180D hosp. mortality
indicators: 42% of attribution
BMI
Platelets
Pulse Hrt Rate
Temperature
BUN
Eosinophils
Chloride
Creatinine
Sodium
RDW
MCV
WBC
Calcium
Resp Rate
Albumin
Monocytes
MCHC
Systolic BP
Lymphocytes
Neutrophil
Basophils
Diastolic BP
O2
MCH
ALP
AST
HCT
Potassium
CO2
Bilirubin, Total
HGB
RBC
Glucose
Protein, Total
ALT
0
1
2
3
4
5
6
Share of total LIME attribution (%)

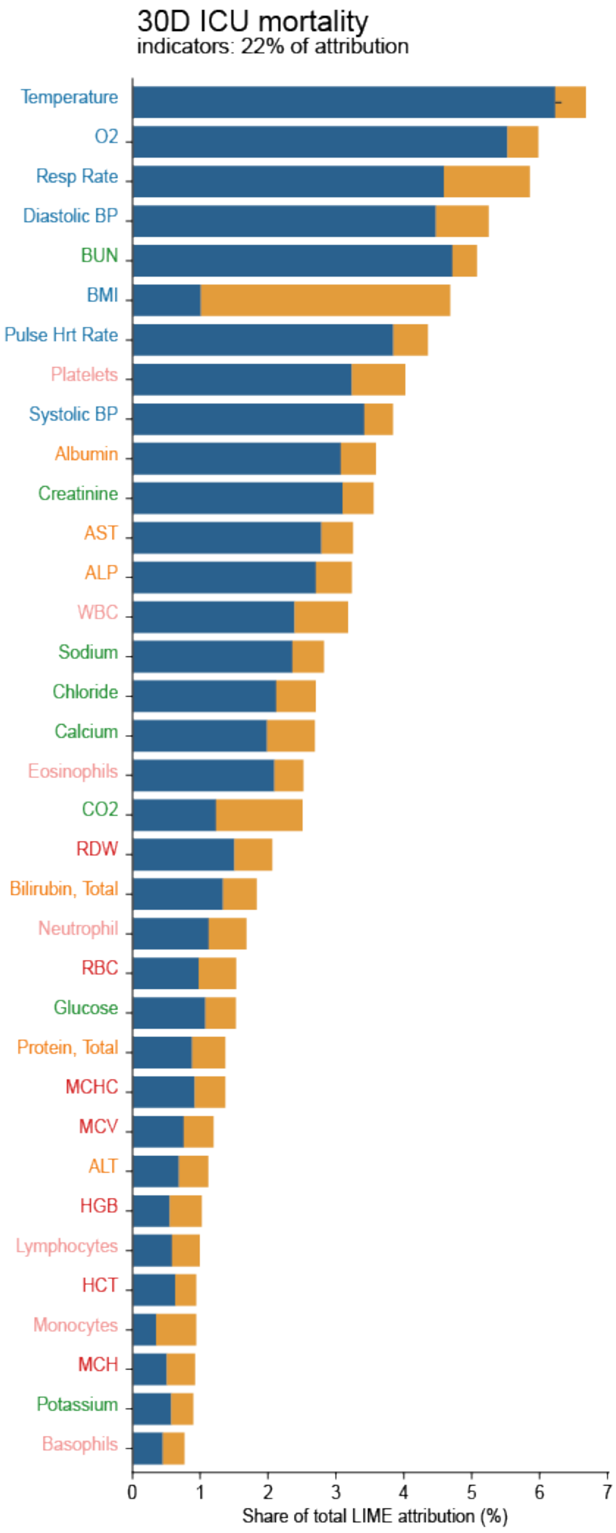

30D ICU mortality
indicators: 22% of attribution
Temperature
O2
Resp Rate
Diastolic BP
BUN
BMI
Pulse Hrt Rate
Platelets
Systolic BP
Albumin
Creatinine
AST
ALP
WBC
Sodium
Chloride
Calcium
Eosinophils
CO2
RDW
Bilirubin, Total
Neutrophil
RBC
Glucose
Protein, Total
MCHC
MCV
ALT
HGB
Lymphocytes
HCT
Monocytes
MCH
Potassium
Basophils
0
1
2
3
4
5
6
7
Share of total LIME attribution (%)

b.

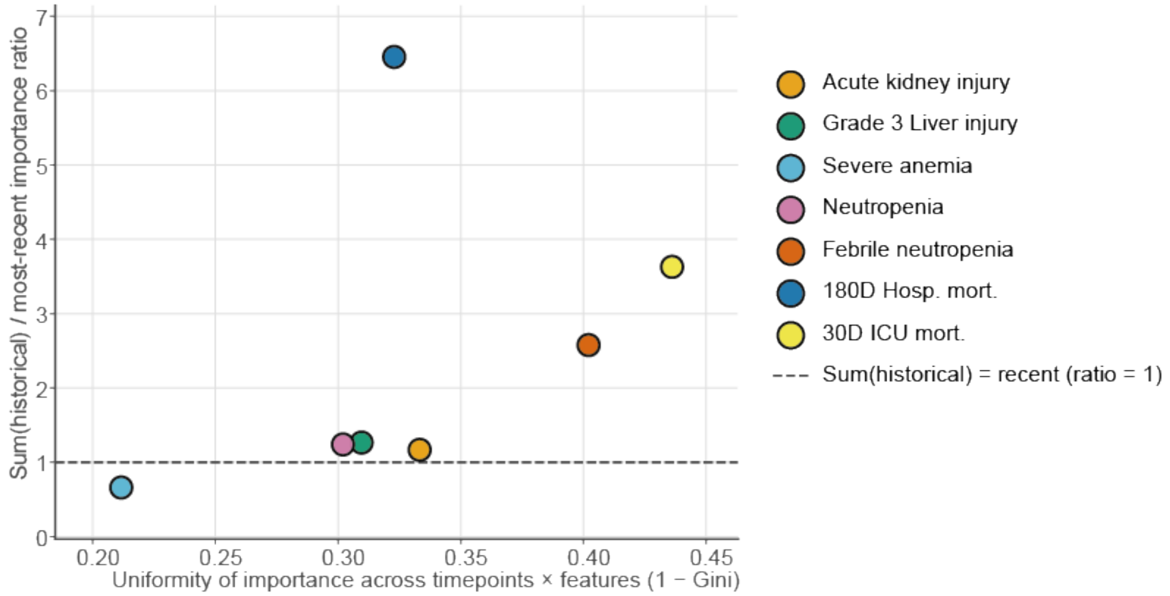

Sum(historical) / most-recent importance ratio
0
1
2
3
4
5
6
7
0.20
0.25
0.30
0.35
0.40
0.45
Uniformity of importance across timepoints × features (1 − Gini)
Acute kidney injury
Grade 3 Liver injury
Severe anemia
Neutropenia
Febrile neutropenia
180D Hosp. mort.
30D ICU mort.
Sum(historical) = recent (ratio = 1)

c.

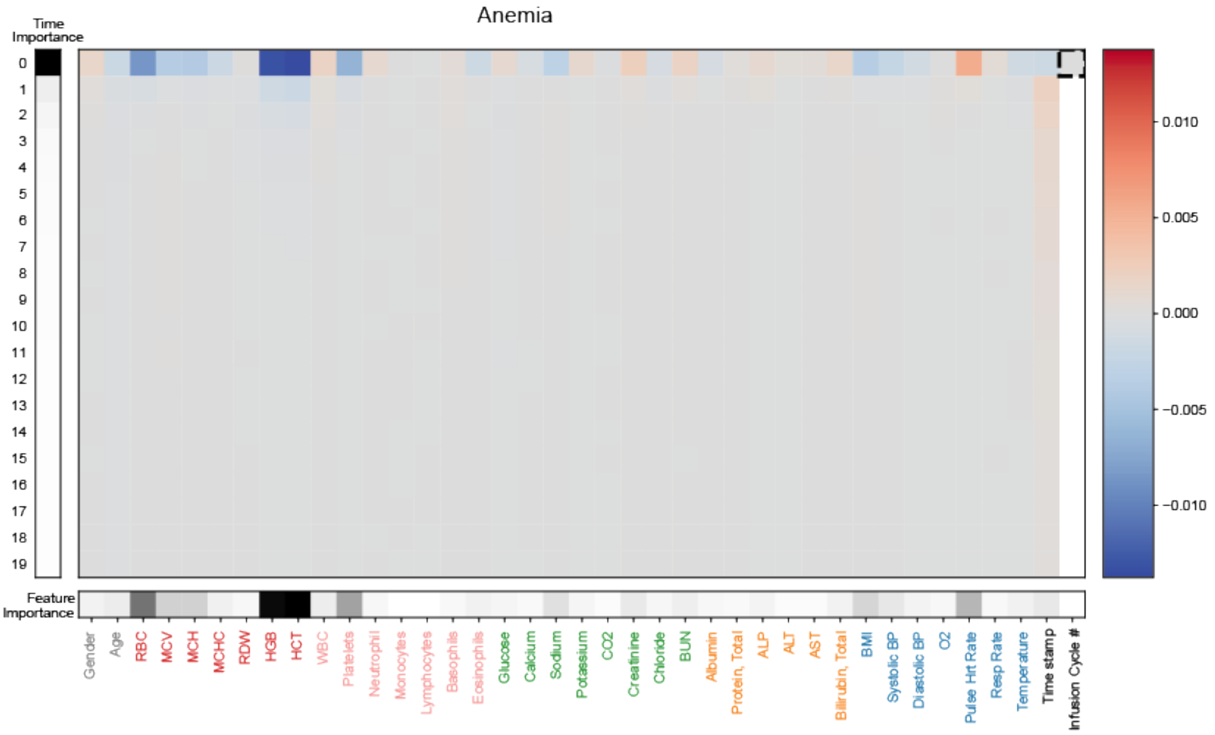

Anemia
Time Importance
0.010
0.005
0.000
−0.005
−0.010
Feature Importance
Gender
Age
RBC
MCV
MCH
MCHC
RDW
HGB
HCT
WBC
Platelets
Neutrophil
Monocytes
Lymphocytes
Basophils
Eosinophils
Glucose
Calcium
Sodium
Potassium
CO2
Creatinine
Chloride
BUN
Albumin
Protein, Total
ALP
ALT
AST
Bilirubin, Total
BMI
Systolic BP
Diastolic BP
O2
Pulse Hrt Rate
Resp Rate
Temperature
Time stamp
Infusion Cycle #

d.

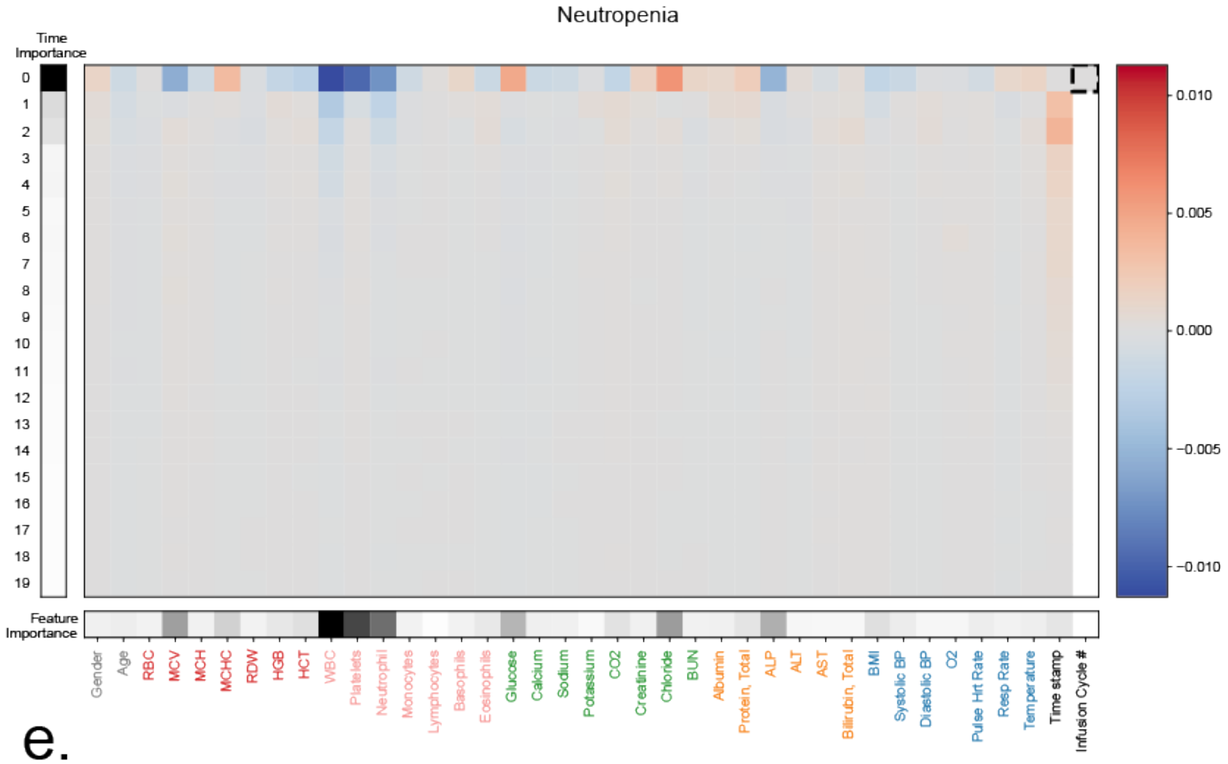
Neutropenia
Time Importance
0 1 2 3 4 5 6 7 8 9 10 11 12 13 14 15 16 17 18 19
0.010
0.005
0.000
−0.005
−0.010
Feature Importance
Gender Age RBC MCV MCH MCHC RDW HGB HCT WBC Platelets Neutrophil Monocytes Lymphocytes Basophils Eosinophils Glucose Calcium Sodium Potassium CO2 Creatinine Chloride BUN Albumin Protein, Total ALP ALT AST Bilirubin, Total BMI Systolic BP Diastolic BP O2 Pulse Hrt Rate Resp Rate Temperature Time stamp Infusion Cycle #

e.

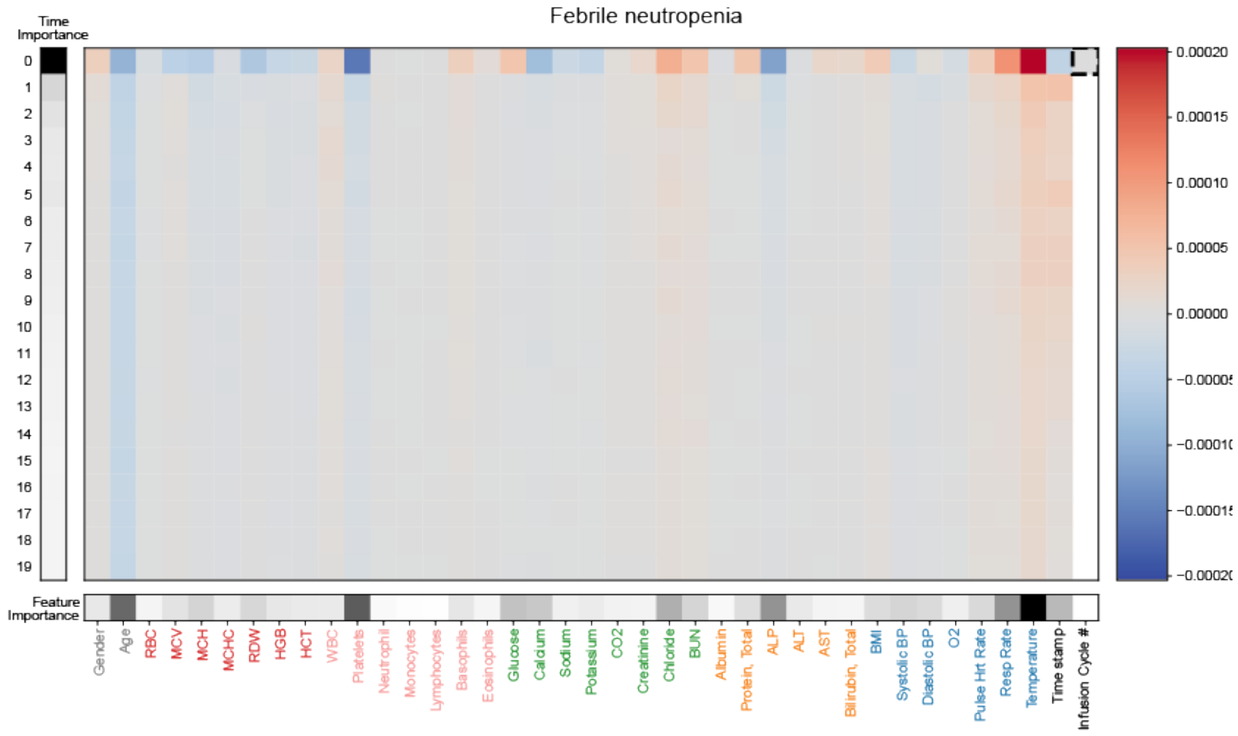
Febrile neutropenia
Time Importance
0 1 2 3 4 5 6 7 8 9 10 11 12 13 14 15 16 17 18 19
0.00020
0.00015
0.00010
0.00005
0.00000
Feature Importance
Gender Age RBC MCV MCH MCHC RDW HGB HCT WBC Platelets Neutrophil Monocytes Lymphocytes Basophils Eosinophils Glucose Calcium Sodium Potassium CO2 Creatinine Chloride BUN Albumin Protein, Total ALP ALT AST Bilirubin, Total BMI Systolic BP Diastolic BP O2 Pulse Hrt Rate Resp Rate Temperature Time stamp Infusion Cycle #

f.

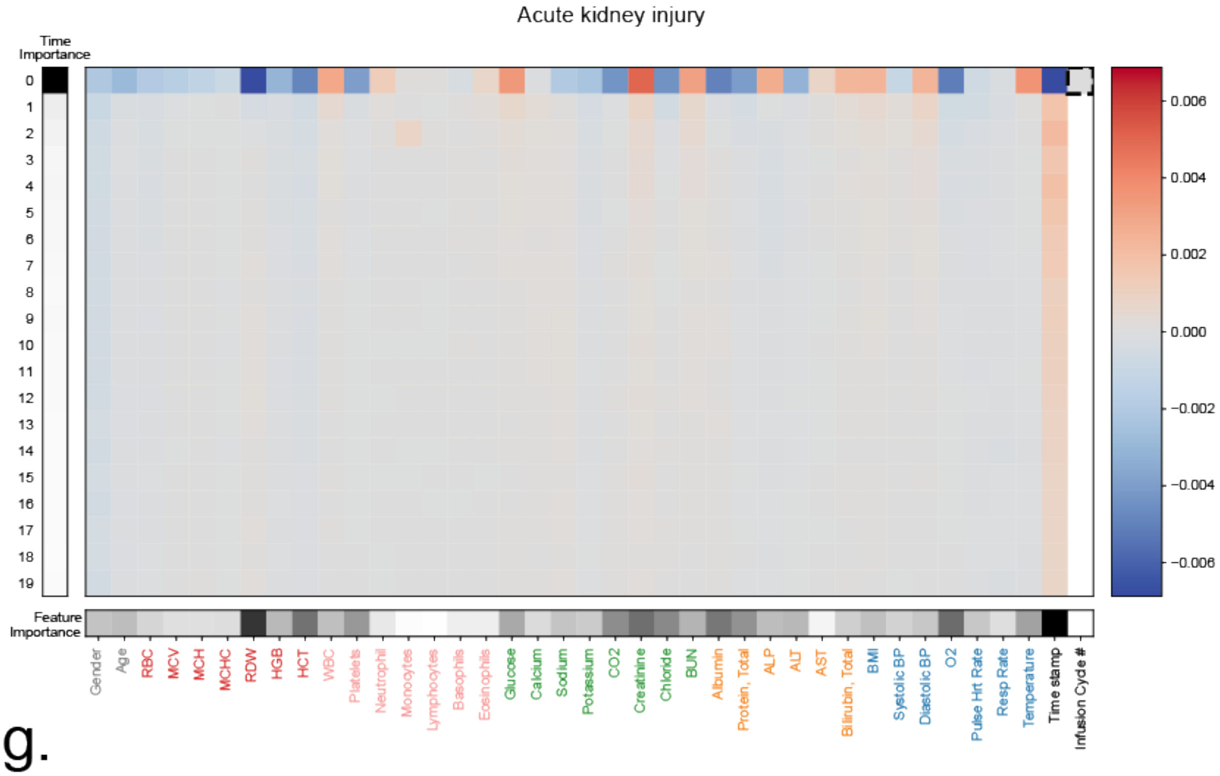
Acute kidney injury
Time Importance
Feature Importance
Gender
Age
RBC
MCV
MCH
MCHC
RDW
HGB
HCT
WBC
Platelets
Neutrophil
Monocytes
Lymphocytes
Basophils
Eosinophils
Glucose
Calcium
Sodium
Potassium
CO2
Creatinine
Chloride
BUN
Albumin
Protein, Total
ALP
ALT
AST
Bilirubin, Total
BMI
Systolic BP
Diastolic BP
O2
Pulse Hrt Rate
Resp Rate
Temperature
Time stamp
Infusion Cycle #
0.006
0.004
0.002
0.000
−0.002
−0.004
−0.006

g.

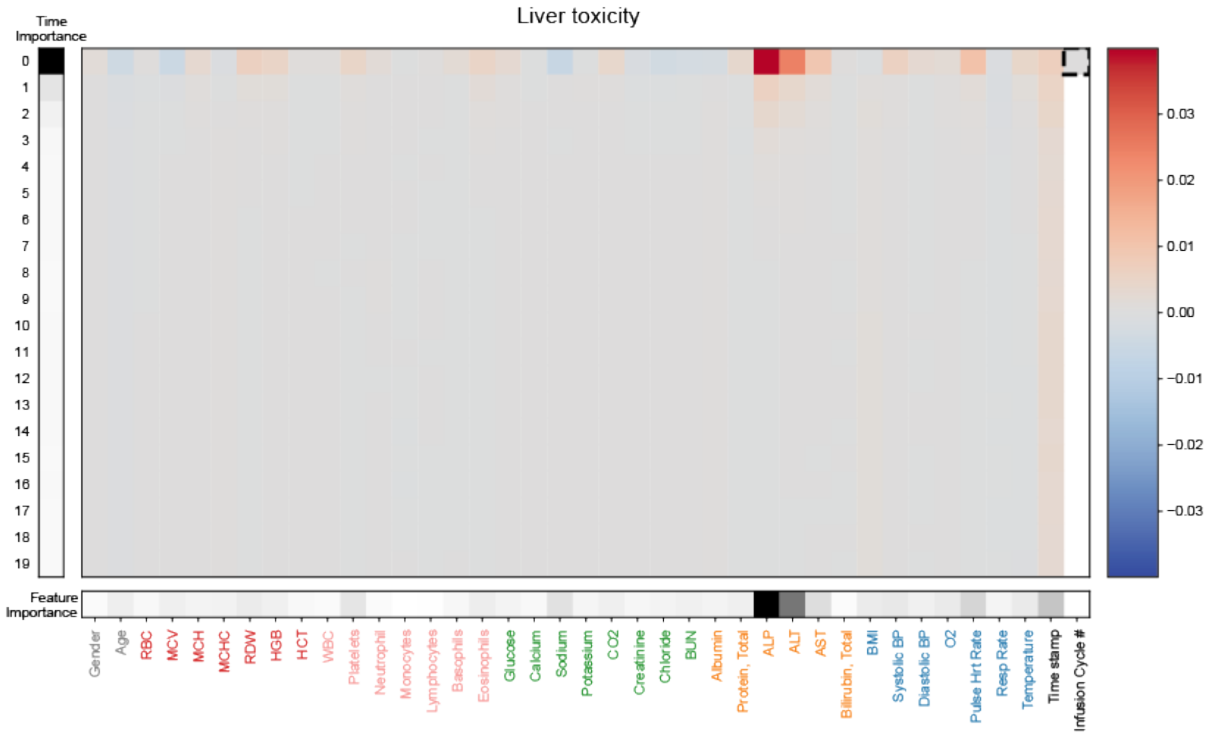
Liver toxicity
Time Importance
Feature Importance
Gender
Age
RBC
MCV
MCH
MCHC
RDW
HGB
HCT
WBC
Platelets
Neutrophil
Monocytes
Lymphocytes
Basophils
Eosinophils
Glucose
Calcium
Sodium
Potassium
CO2
Creatinine
Chloride
BUN
Albumin
Protein, Total
ALP
ALT
AST
Bilirubin, Total
BMI
Systolic BP
Diastolic BP
O2
Pulse Hrt Rate
Resp Rate
Temperature
Time stamp
Infusion Cycle #
0.03
0.02
0.01
0.00
−0.01
−0.02
−0.03

h.

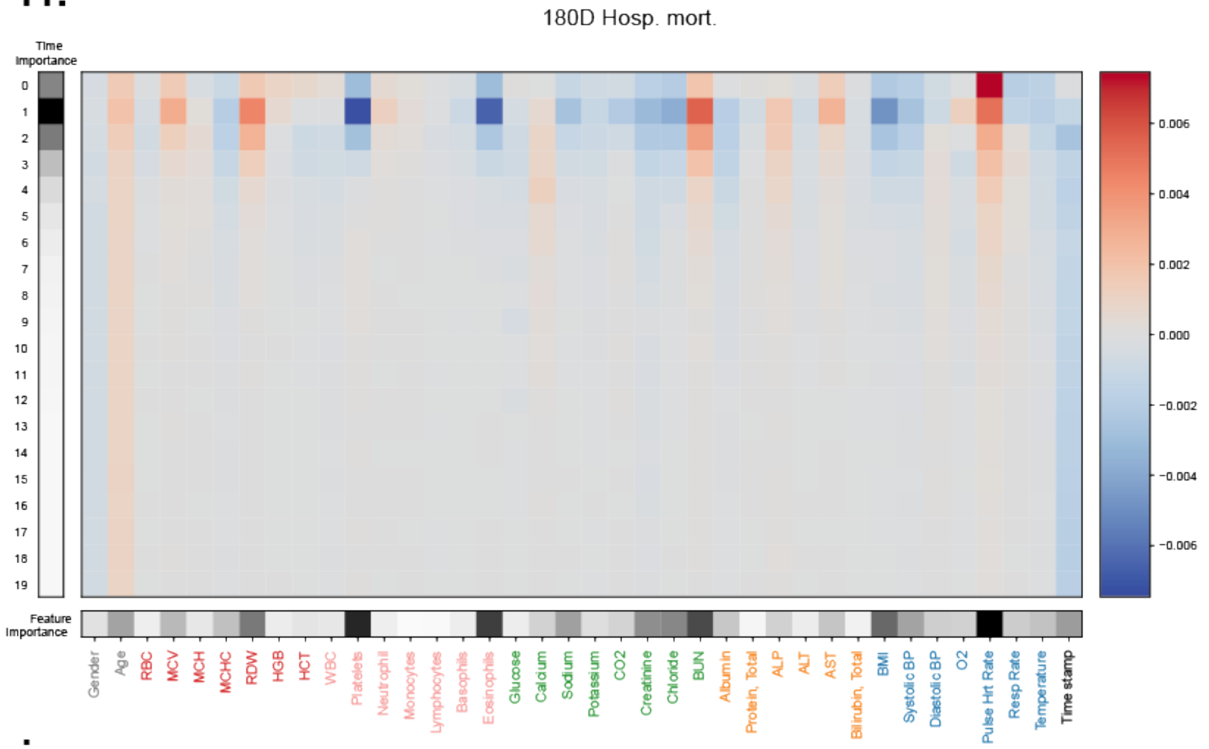

i.

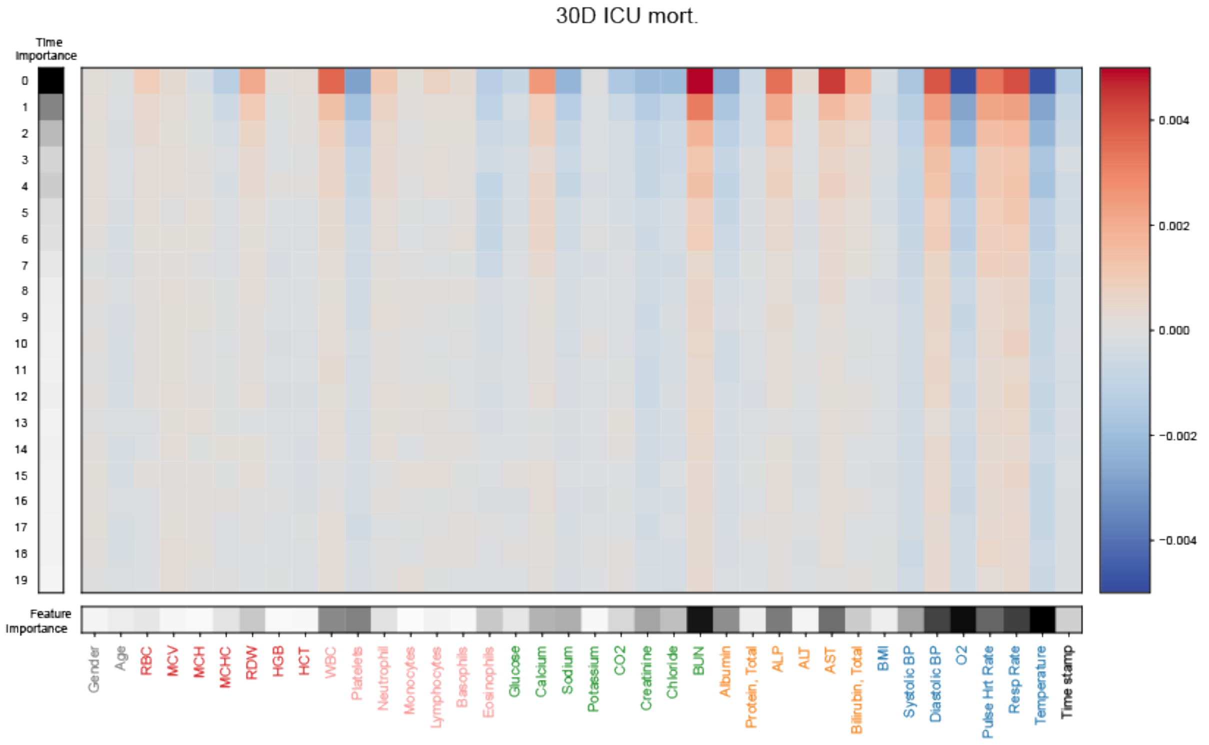

**Supplemental Figure 6. Additional feature importance analyses.** (a) Temporal importance analyses and (b) per-task feature importance analyses, with signed LIME values in the main panel and averaged-then-absolute importance over time and features calculated at the left and bottom. see Methods for methodology.

a.

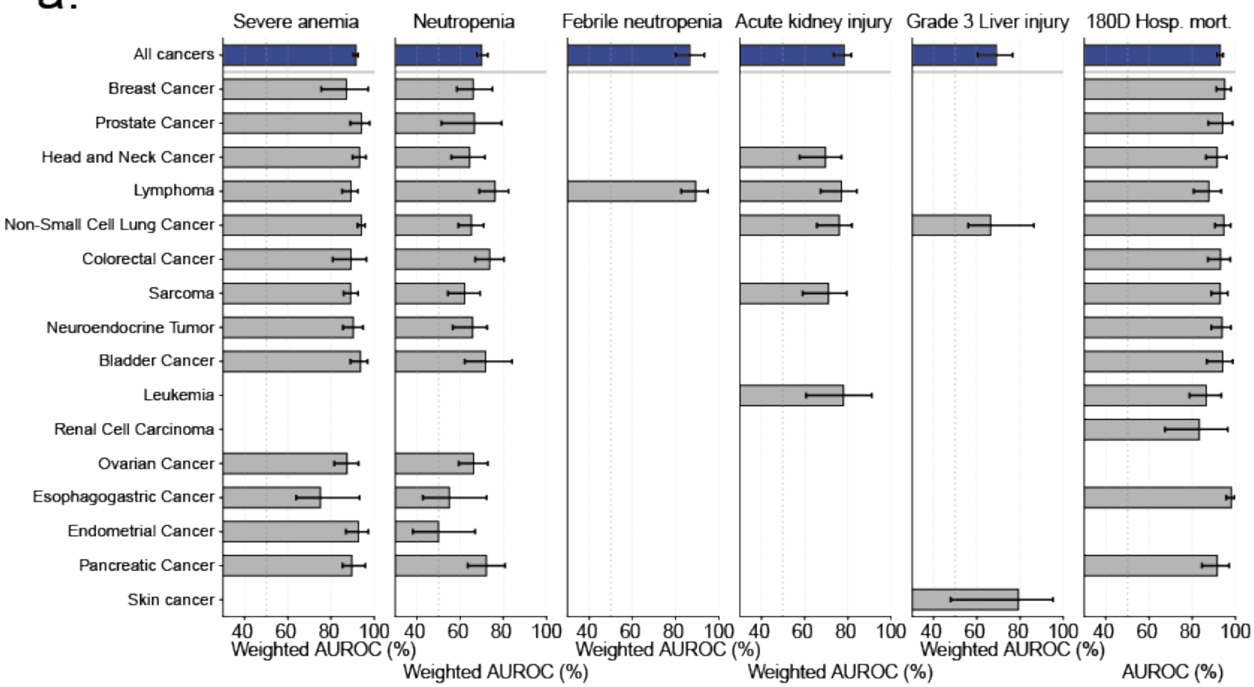

b.

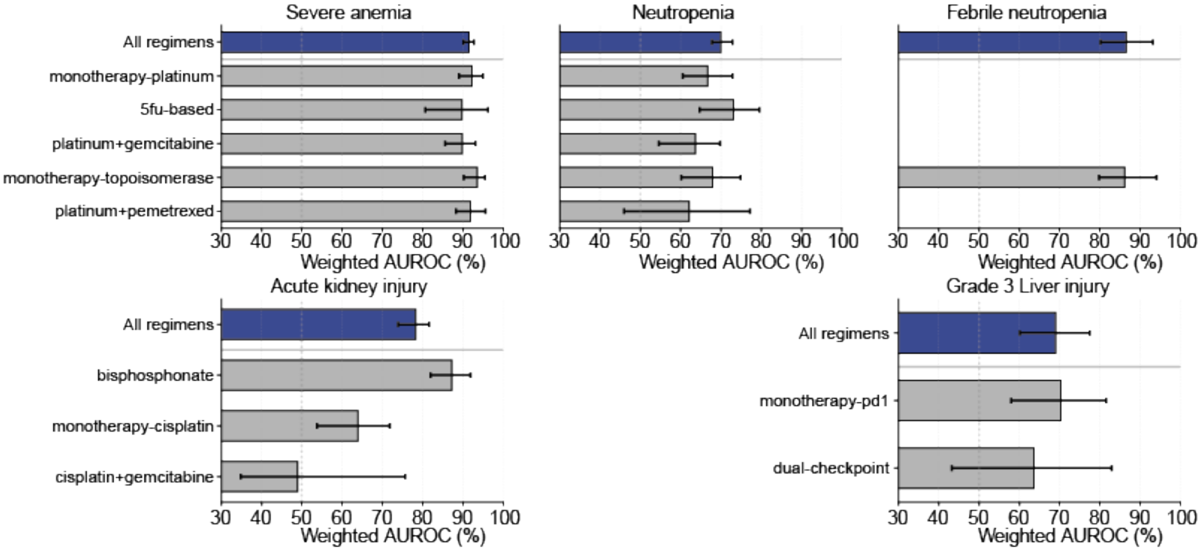

**Supplemental Figure 7. SPARC performance stratified by (a) cancer types and (b) treatment regimen.** See Tables S6 and S7 for detailed numbers visualized on this figure.

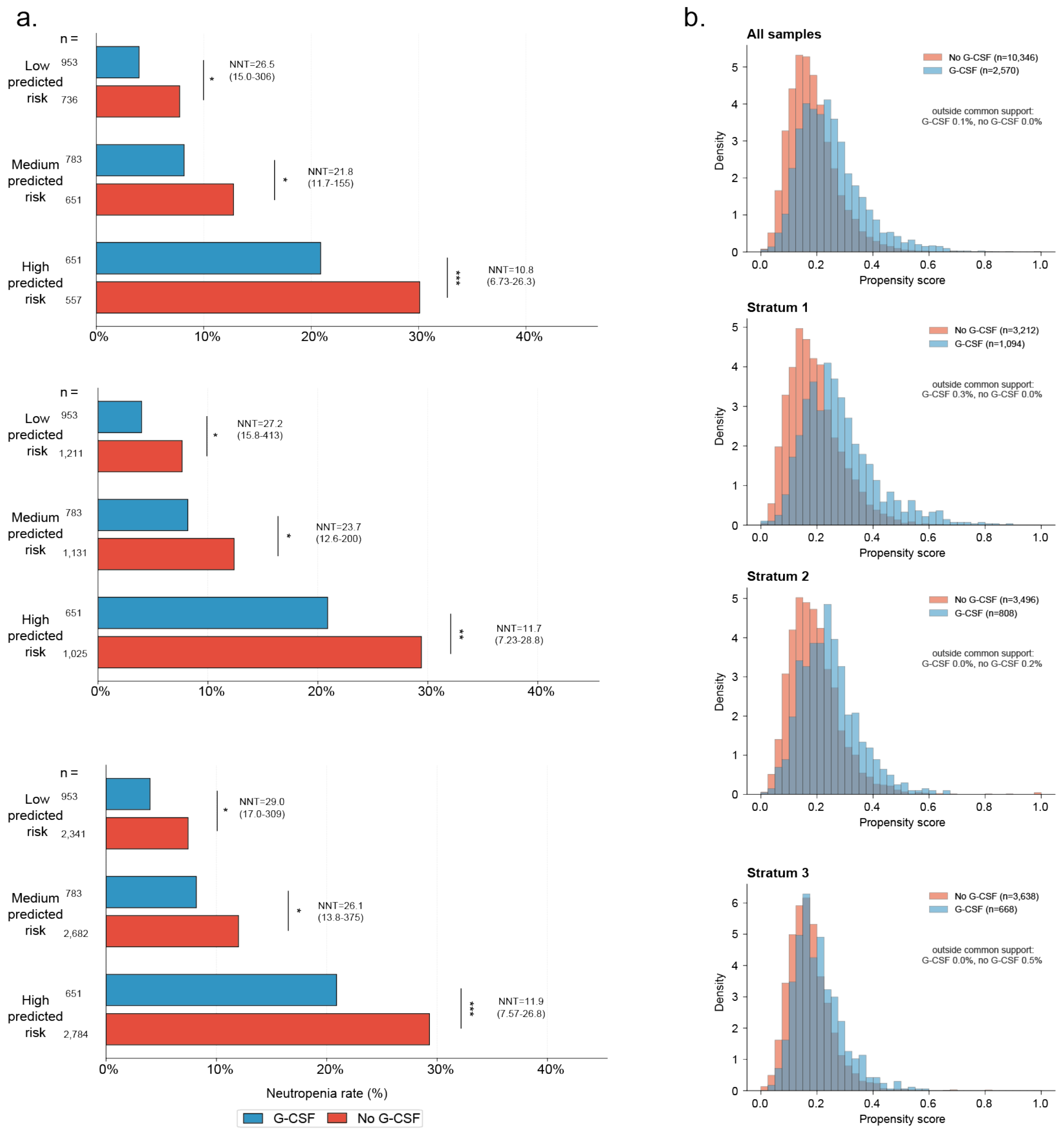

**Supplementary Figure 8. Propensity score matching and positivity for the G-CSF analysis.** (a) Neutropenia rates with and without G-CSF within tertiles of SPARC-predicted risk after propensity score matching with 1:1 (top), 1:2 (middle) and 1:10 (bottom) controls per G-CSF-treated sample. Matching was conducted by nearest neighbour on the logit propensity score with a caliper of 0.2 SD, with replacement. Numbers next to the bars are the G-CSF number of samples. NNT, number needed to treat (1/absolute risk reduction); 95% CIs and p-values from a patient-level bootstrap (2,000 resamples); *p < 0.05, **p < 0.01, ***p < 0.001. (b) Distributions of propensity scores (probability of receiving G-CSF, as used for weighting) for G-CSF-treated and untreated infusions, overall and within each risk tertile (strata 1–3, low to high predicted risk).

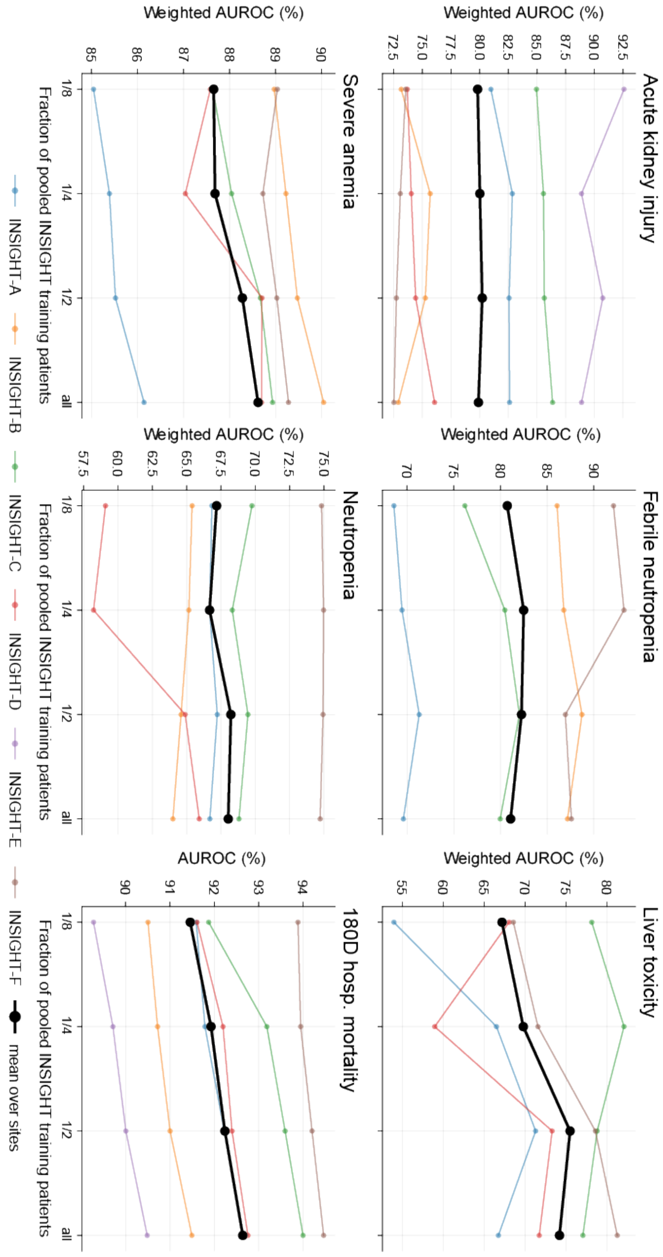

**Supplementary Figure 9. SPARC performance saturation analysis on INSIGHT data.** We randomly titrated the pooled training data to its fractions (x-axis) and compared the performance (y-axis) of the trained models.
